# Multimodal Foundation Models for Medical Imaging - A Systematic Review and Implementation Guidelines

**DOI:** 10.1101/2024.10.23.24316003

**Authors:** Shih-Cheng Huang, Malte Jensen, Serena Yeung-Levy, Matthew P. Lungren, Hoifung Poon, Akshay S Chaudhari

## Abstract

Advancements in artificial intelligence (AI) offer promising solutions for enhancing clinical workflows and patient care, potentially revolutionizing healthcare delivery. However, the traditional paradigm of AI integration in healthcare is limited by models that rely on single input modalities during training and require extensive labeled data, failing to capture the multimodal nature of medical practice. Multimodal foundation models, particularly Large Vision Language Models (VLMs), have the potential to overcome these limitations by processing diverse data types and learning from large-scale unlabeled datasets or natural pairs of different modalities, thereby significantly contributing to the development of more robust and versatile AI systems in healthcare. In this review, we establish a unified terminology for multimodal foundation models for medical imaging applications and provide a systematic analysis of papers published between 2012 and 2024. In total, we screened 1,144 papers from medical and AI domains and extracted data from 97 included studies. Our comprehensive effort aggregates the collective knowledge of prior work, evaluates the current state of multimodal AI in healthcare, and delineates both prevailing limitations and potential growth areas. We provide implementation guidelines and actionable recommendations for various stakeholders, including model developers, clinicians, policymakers, and dataset curators.

## 1. Introduction

Artificial Intelligence (AI) in healthcare presents significant opportunities to transform clinical workflows and patient care, ultimately improving patient outcomes. Despite numerous attempts to leverage AI models for healthcare, a significant gap remains between AI’s potential and its currently usefulness in clinical practice^1–4^. For instance, most contemporary healthcare AI models are constrained by a reliance on single input modalities during training, failing to capture the multimodal nature of medical practice^5^. This contrasts with real-world clinical practice, where physicians rely on diverse data sources to form a holistic view of patient health^6,7^. Moreover, the prevalent use of supervised learning requires extensive, clinical specialist-curated labels, a process that is neither scalable nor cost-effective, leading to models that excel in narrow tasks without broader applicability^8^. To bridge this gap and fully realize AI’s potential in healthcare, a paradigm shift is imperative: there is a need for AI models that are capable of processing multimodal inputs during training and learning from vast amounts of unlabeled data or natural pairs of different modalities, such as medical images and their corresponding reports. These approaches can enhance the performance and usefulness of AI in medical settings, heralding a new era of AI-driven healthcare innovations.

Recently, the AI field has witnessed a leap in capabilities driven by advanced Foundation Models^9^ such as GPT^10^ and Llama^11^. Unlike previous generations of specialized models, these Foundation Models can perform a wide variety of tasks using a single model trained on vast amounts of data^9^, typically through a training strategy called self-supervised learning (see Terminologies and Strategies section). Subsequently, these models exhibit emergent capabilities on tasks for which they were not explicitly trained^9^. Examples of emergent properties include zero-shot learning, where a model can identify e.g. a disease it was not explicitly trained to classify. While many pioneering Foundation Models are trained with text, which offers a direct semantic interface for humans to intuitively interact with the Foundation Models, these models are not restricted to text only. In fact, several recent research efforts have focused on multimodal foundation models that can integrate additional modalities, such as GPT-4V(ision)^12^, LLaVA^13^, and Gemini^14^. Multimodal models have a large potential for clinical use since patient data often include several modalities.

While these models show great promise, they are still in their nascent stages in healthcare. The pathway to developing clinically useful tools remains challenging, requiring advancements in accuracy, safety, and workflow integration. The potential to effect positive changes in healthcare and improve patient outcomes hinges not only on the abilities of model developers but also requires a concerted effort from clinicians, policymakers, and dataset curators^4^. Clinicians play a pivotal role in identifying genuine clinical needs and determining the essential modalities for specific medical tasks. Policymakers are instrumental in updating their policies to consider the nuances of multimodal foundation models and striking a balance between streamlining the approval process and keeping a high standard for safety. Dataset curators must prioritize the collection of diverse, representative and multimodal data while maintaining high quality and clinical relevance. Interdisciplinary collaboration, guided by a shared language and understanding of these complex issues, is crucial to address current challenges and guide future model development.

The objective of this review is threefold: i) to establish and unify the terminology critical to the intersection of AI and healthcare, with an emphasis on multimodal SSL training (see Terminologies and Strategies); ii) to conduct a systematic review of the new field of multimodal Foundation Models for medical imaging applications, extracting key insights and evaluating their current state (see Results); and iii) to highlight the prevailing limitations and actionable future strategies for a broad array of stakeholders, including model developers, clinicians, policymakers, and dataset curators (See Discussion and Guidelines). We focused on multimodality involving medical images, such as radiology image and pathology slides, since medical imaging is an essential part of the diagnostic and treatment workflow across various medical specialties. Although recent trends in medical imaging AI literature increasingly focus on utilizing multimodal foundation models (see Figure. 1), with a handful of narrative reviews available^15–18^, there are currently no systematic reviews. By adhering to the PRISMA^19^ guidelines, we methodically gather and consolidate the latest contributions of multimodal Foundation Models for medical imaging applications, providing a comprehensive snapshot of the existing landscape. In total, we screened 1,144 papers and extracted data from 97 papers for this systematic review. Our investigation identifies both challenges and their potential solutions for the deployment of multimodal Foundation Models, with a focus on advancing their usefulness in healthcare.

**Figure 1:**
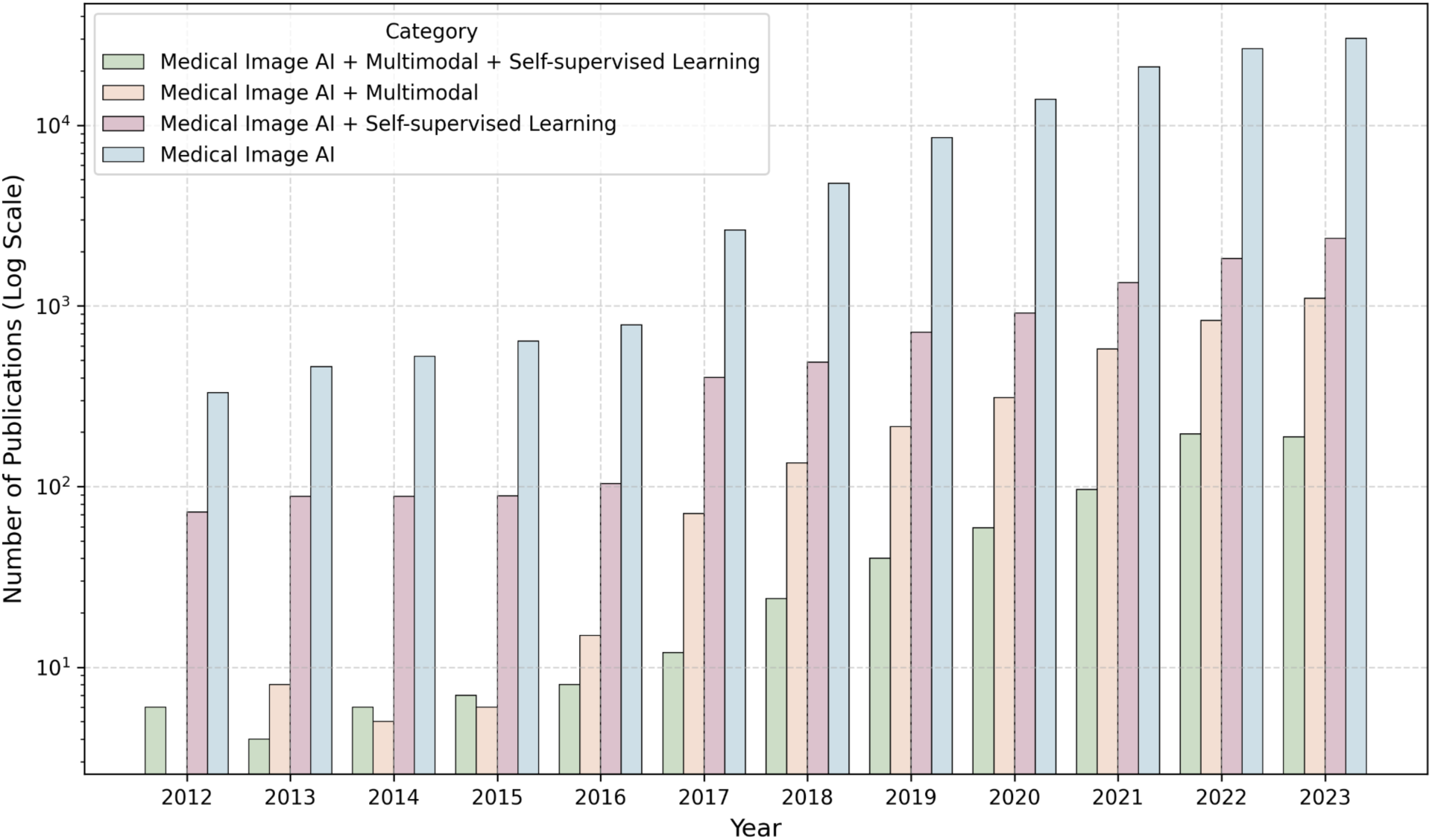
Timeline showing growth in publications on deep learning for medical imaging, based on search criteria applied to PubMed and Scopus. The figure illustrates that multimodal self-supervised learning represents a small but rapidly growing subset of medical deep learning literature. Publication counts were aggregated using keyword groups. For example, “Medical AI” combines the “Deep Learning” and “Medical Imaging” groups, while “Medical AI + Self-supervised Learning” includes the prior two groups plus the “Self-supervised Learning” group. Specific keywords for each group are detailed in the Methodology section and Appendix. Y-axis is in log scale.

## 2. Terminology and strategies

The development of foundation models typically involves a two-stage training process: pretraining and fine-tuning. During the pretraining stage, the vast majority of foundation models employ self-supervised strategies - a process that utilizes large volumes of unlabeled or naturally paired data to learn general, transferable, and label-efficient representations. Subsequently, in the fine-tuning stage, the pretrained model is adapted to specific downstream tasks. Owing to the knowledge acquired during pretraining, fine-tuning often necessitate minimal labeled data, and in some cases, can be accomplished without task-specific labels.

In this section, we provide definitions for different categories of multimodal self-supervised pretraining strategies: contrastive, self-prediction, generative, and generative Vision-Language Models (VLMs) (Figure 2). Additionally, we illustrate various approaches for adapting pretrained models to downstream tasks through fine-tuning (Figure 3).

**Figure 2:**
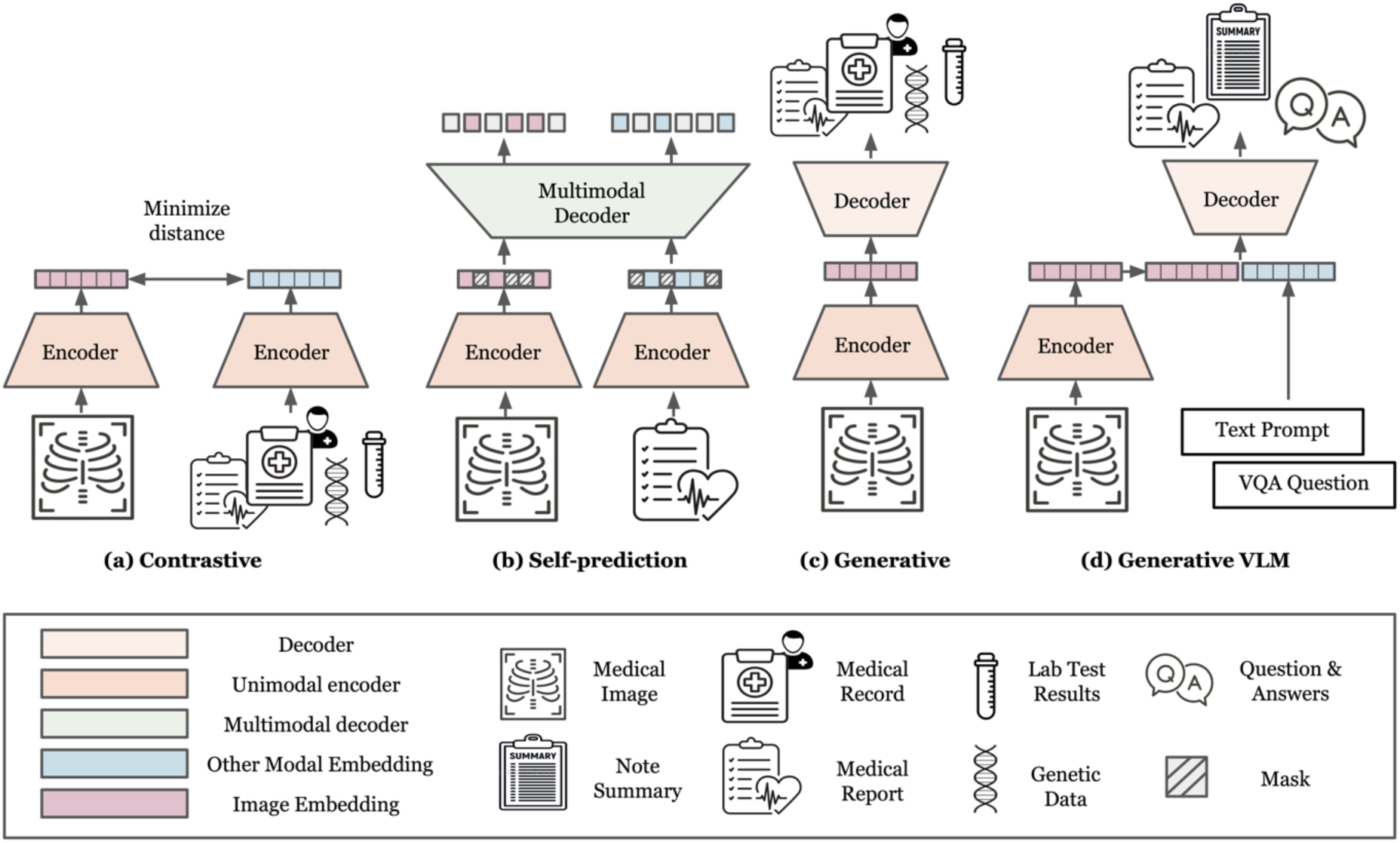
Illustration of multimodal self-supervised learning pretraining strategies. During the pre-training stage of multimodal Foundation Models, one or more of the following self-supervised strategies are typically used: (**a**) Contrastive Learning forms positive pairs between matching data with shared semantic content, e.g. X-ray images and reports for the same medical examination, and minimizes the representational distance in a common latent space of positive samples (**b**) Self-prediction masks out random parts of the inputs and seeks to reconstruct the masked out regions by utilizing complimentary information across the input modalities (**c**) Generative SSL learns the distribution of the training data by generating one or several modalities from another, e.g. generating a report from an X-ray or vice versa (**d**) Generative VLM is a special case of Generative SSL, where an input instruction (“prompt”) can be used to steer the output generated by the model.

**Figure 3:**
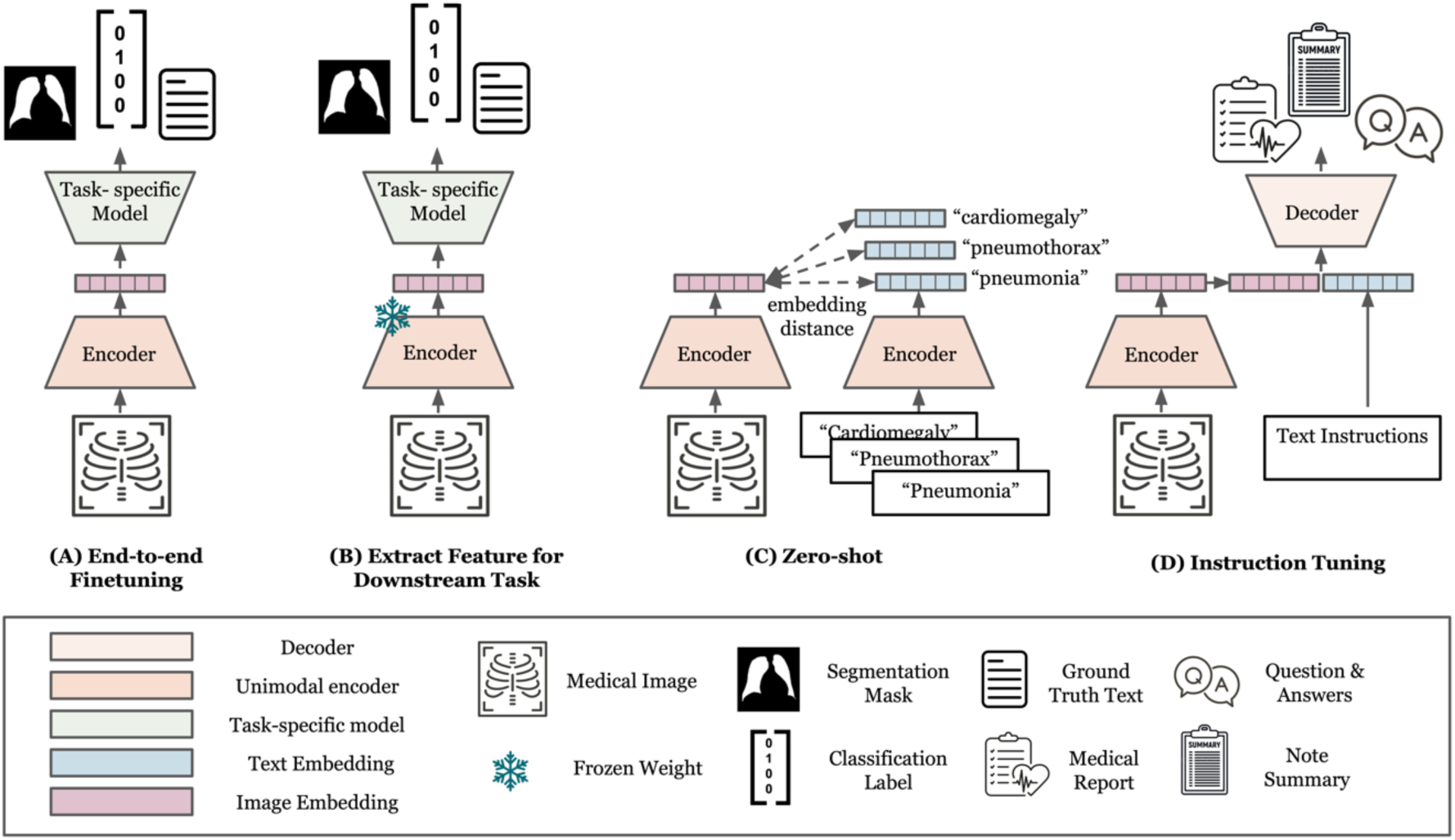
Illustration of strategies for adapting pretrained models to downstream tasks. During the fine-tuning stage of a pretrained multimodal multimodal Foundation Model, one or several of the following strategies can be used to adapt the model to a given downstream task: (**a**) The entire or parts of the pretrained model is finetuned for the downstream task via supervised learning (**b**) The encoder is frozen and used only as a feature extractor, while a task-specific model is trained to utilize these features for the downstream task using supervised learning. (**c**) Pre-trained model embedding text prompts describing potential classes alongside the image and predicting the class whose text embedding is closest to the image embedding in the shared latent space without additional training. (**d**) The model, typically a VLM, is fine-tuned using pairs of instructions and expected outputs for the downstream task.

### 2.1 Contrastive

Contrastive self-supervised learning paradigms presuppose that semantically similar input pairs, termed ‘positive pairs,’ should exhibit closer alignment in feature space compared to disparate inputs, or ‘negative pairs’. Pioneering methodologies, exemplified by SimCLR^20^ and MoCo^21^, predominantly focused on unimodal data—specifically images. The core objective underpinning these models is the dual process of minimizing the distance between embeddings of positive pairs and maximizing that between negative pairs. Multiple approaches can be used to form positive and negative pairs, where the most common are various augmentations of the same inputs to constitute semantically similar positive pairs, while augmentations across distinct inputs from negative pairs.

Progressing beyond unimodal frameworks, Contrastive Language-Image Pre-Training (CLIP^22^) integrates contrastive learning across image and textual domains. The key difference to its unimodal predecessors is that CLIP delineates positive pairs as images and their corresponding captions, seeking to co-locate image and textual descriptions within a unified multimodal representation space. This approach has paved the way for further explorations into multimodal contrastive learning, yielding diverse strategies for generating localized positive pairs between images and text, hence discovering more fine grained image-text associations^23–25^. Notably, the scope of modalities encompassed by recent advancements is not confined to images and text but extends to other modalities such as acoustic signals, electronic health records, or sensor data, provided the paired modalities convey shared semantic content.

### 2.2 Self-prediction

Self-prediction SSL involves the process of masking parts of the input data and subsequently attempting to reconstruct the original, unmodified input (Figure 2B). Self-prediction first emerged in the natural language processing (NLP) domain, where state-of-the-art models were initially trained through a process called Masked Language Modeling (MLM), which involves predicting the masked words from a sentence^26^. Inspired by this success in NLP, initial experiments in computer vision also adopted this method by obscuring or altering random patches of images and training Convolutional Neural Networks (CNNs) to fill in the gaps as a pre-training method^27,28^. More recently, the advent of Vision Transformers (ViT)^29^ enabled adopting transformer-based architectures that have shown considerable success in NLP research. Techniques such as BERT Pre-Training of Image Transformers (BEiT)^30^ and Masked Autoencoders (MAE)^31^ that utilize self-prediction in conjunction with ViT have demonstrated superior performance after fine-tuning on various natural image benchmarks compared to their CNN based predecessors.

In a multi-modal setting, one or several of the modalities can be masked out, before the reconstruction step^32^. This approach allows the model to leverage the complementary information across multiple modalities when reconstructing the masked segments, thereby facilitating enhanced understanding of the complex associations between the modalities. Often corresponding text and images are used, such as X-rays and radiology reports, where e.g. parts of the image or text are masked out, and the information from both modalities is used concurrently to reconstruct the input^33^. However, self-prediction may be extended to any other modalities, such as genetics, blood panels, sensor data, or other medical data.

### 2.3 Generative

Generative models have been developed to learn the distribution of training data, which allows them to either reconstruct original inputs or generate new, synthetic data. Unlike self-prediction SSL methods that focus solely on masking parts of the input and uses the rest of the unmodified input to guide the reconstruction process, generative SSL methods modify the entire input and aim to reconstruct it as a whole. Hence, while self-prediction can only fill in removed information, generative approaches can generate new data.

Pioneering work on generative models utilized autoencoders^34^. Here, an encoder transforms high-dimensional inputs into a lower-dimensional compressed version (latent representation), followed by a decoder that uses the latent representation to reconstruct the original high-dimensional input. In a multi-modal setting, an encoder would take one modality as input and the decoder would generate another modality^35^ (Figure 2C). For instance, an encoder can take in medical images as inputs, and the decoder’s task is to generate the corresponding reports^36,37^.

Following autoencoders, Generative Adversarial Networks (GANs)^38^ and diffusion models^39^ have achieved notable success and popularity, particularly for image generation tasks. GANs use a generative model to generate a high quality output followed by a discriminator network trying to distinguish whether the generated output hails from the original data distribution or is a synthetic output. Diffusion models are trained by progressively adding small amounts of artificial noise to an input, with the goal of learning to reverse this process and iteratively denoising the input with the added noise. Successively adding even small amounts of noise to an input eventually fully converts it to noise. This can be leveraged during inference since the model has been trained to denoise inputs. At test time, pure noise can be progressively denoised to end up with an input that resembles the original data distribution.

In multimodal settings, both GANs and diffusion models can incorporate other modalities to condition the generation process. A popular approach is using text prompts to guide the generation. For instance, instead of generating random medical images, these models can be prompted to generate specific types of medical images (e.g., chest X-rays with abnormalities) by incorporating text embeddings from clinical descriptions into the generation process.

### 2.4 Generative VLM

More recently, a new type of multimodal generative model has emerged as a popular way to train foundation models^40^. Here, the encoder takes in an image and an instruction text prompt, and the decoder generates a desired output such as a summary or a detailed description of part of the image (Figure 2D)^13,41^. Typically, this type of generative models can leverage pretrained large language models (LLMs) for both text encoding and decoding which already possess rich semantic understanding, enabling an intuitive input and output language interface for the user. While the majority of these types of Generative VLMs utilize only text and images, these models may also incorporate other modalities such as genetic data, wearable sensors, and other medical data.

### 2.5 Combined Pretraining Approaches

While we have distilled the most common multi-modal pretraining approaches into distinct major categories above, many recent studies combine multiple pretraining approaches, for potentially enriching the model’s pretraining phase by allowing it to leverage the benefits of each approach. The combination of multiple approaches is often achieved by directly optimizing the loss functions of each method or weighting sum of each of the losses. This amalgamation has been empirically demonstrated to enhance performance across various downstream tasks, surpassing models pretrained with a singular pretraining strategy.

An illustrative example of such a combined approach is the Contrastive Captioner (CoCa) model^42^. CoCa integrates two pretraining strategies: generative pretraining, where the model learns to generate text descriptions (captions) for given images using a VLM decoder, and contrastive learning, where it learns to match images with their corresponding text descriptions using CLIP. By combining these strategies, CoCa learns both to describe images in detail and to understand the relationship between images and text at a broader level. This dual approach allows the model to develop a more comprehensive understanding of the connection between visual and textual information.

#### Adapting to Downstream Tasks (Fine-tuning)

Following the label-free pretraining approaches described above, models are typically adapted to specific downstream tasks using labeled data (Figure 3). A common method for doing so involves appending a task-specific head to the pretrained image encoder and fine-tuning the model with a conventional supervised learning regime. This process can be performed in two distinct manners: firstly, by training the entire or parts of the image encoder end-to-end with the task-specific head, as depicted in Figure 3a; alternatively, by freezing the encoder and utilizing it solely as a feature extractor for the task-specific head, thereby leaving encoder’s weights unchanged (Figure 3b).

In the absence of labeled training data, models trained with contrastive learning with images and text can be used to perform zero-shot classification — image classification without the need for any additional training data or labels (Figure 3c). The method poses a class label as text statements, e.g. “a CT scan with ascites present” and “a healthy CT scan”, and both text prompts are then embedded as text embeddings. The proximity of the text embeddings with that of the embedding of the original image is then used to decide what prompt best represents the image^22^.

Alternatively, prompting is a versatile strategy for tasks requiring text generation, such as image captioning, summarization or question answering. In this approach, VLMs are given a textual input (the prompt) that instructs them to output the desired output (Figure 3d). However, the effectiveness of prompting can vary significantly based on the model’s initial pretraining objectives, potentially yielding outputs that diverge from expectations. Addressing this challenge, “instruction tuning” has emerged as a novel training paradigm. This method involves further training of models using explicit pairs of instructions and expected answers tailored to specific downstream tasks. Instruction tuning enhances the model’s ability to follow diverse task-specific prompts and generate text outputs more aligned with the intended task^43^. Some studies have also shown that instruction tuning can enable VLMs to perform a wide range of tasks beyond text generation. For instance, the VLM can be used for classification by instruction tuning it to output classification label as text.^44,45^

## Results

Our systematic search initially identified 1,144 studies. After removing duplicates and applying our selection criteria to the titles and abstracts (detailed in the Methods section), 233 studies qualified for full-text evaluation. Ultimately, 97 studies met our eligibility requirements and were selected for detailed systematic review and data extraction. Out of the 97 studies, 48 studies were multimodal between image and non-image modalities, while 49 used multiple imaging modalities. Figure 4 illustrates the study selection and screening process as a flowchart. The extracted data for included studies that combine medical images with non-image modalities are listed in Table 1 and Supplementary Table 1, while the extracted data for studies with image-only multimodality is listed in Table 2. Figure 5 provides a summary of the statistical analysis of the data.

**Figure 4:**
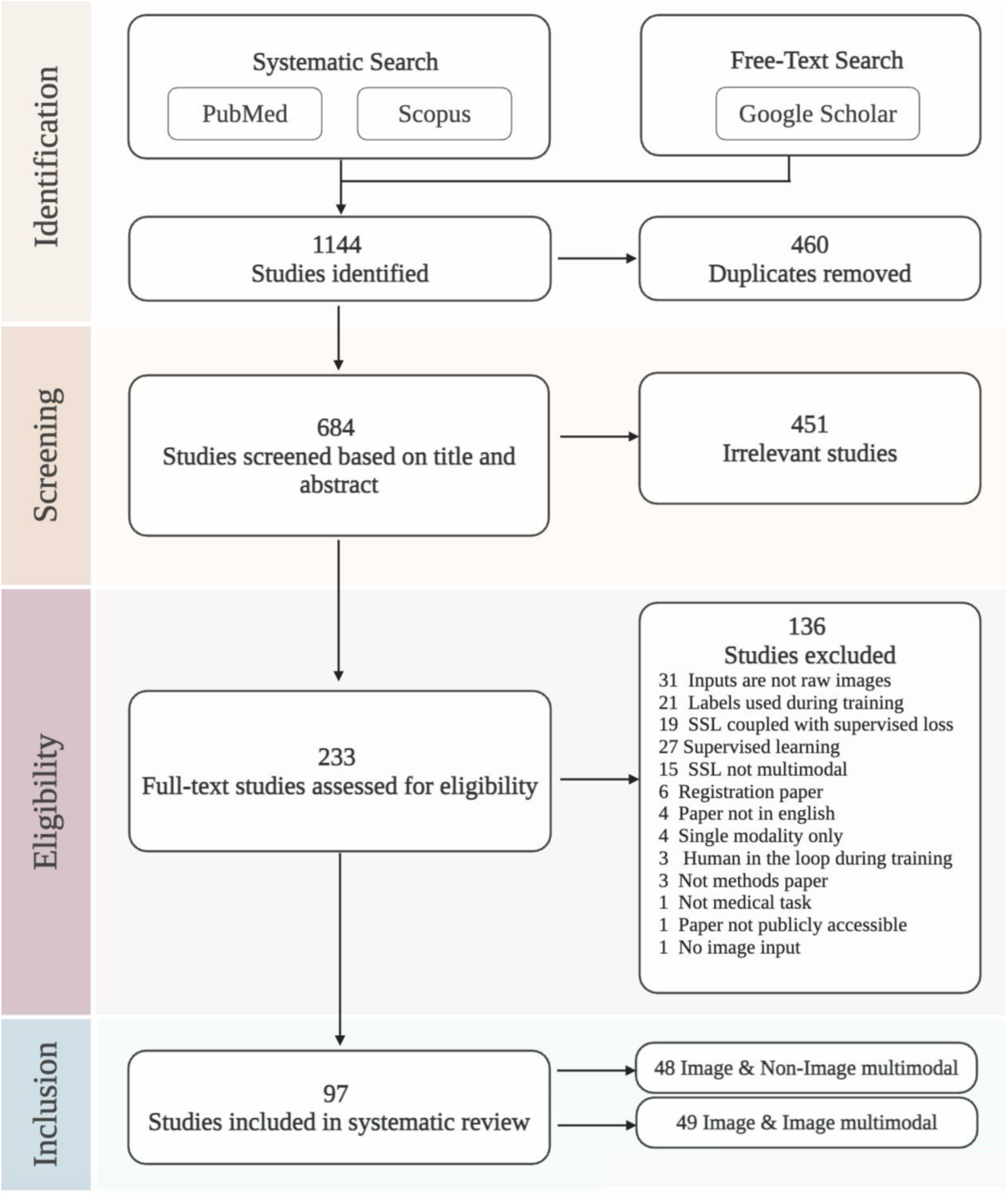
PRISMA flowchart of the study selection process.

**Figure 5:**
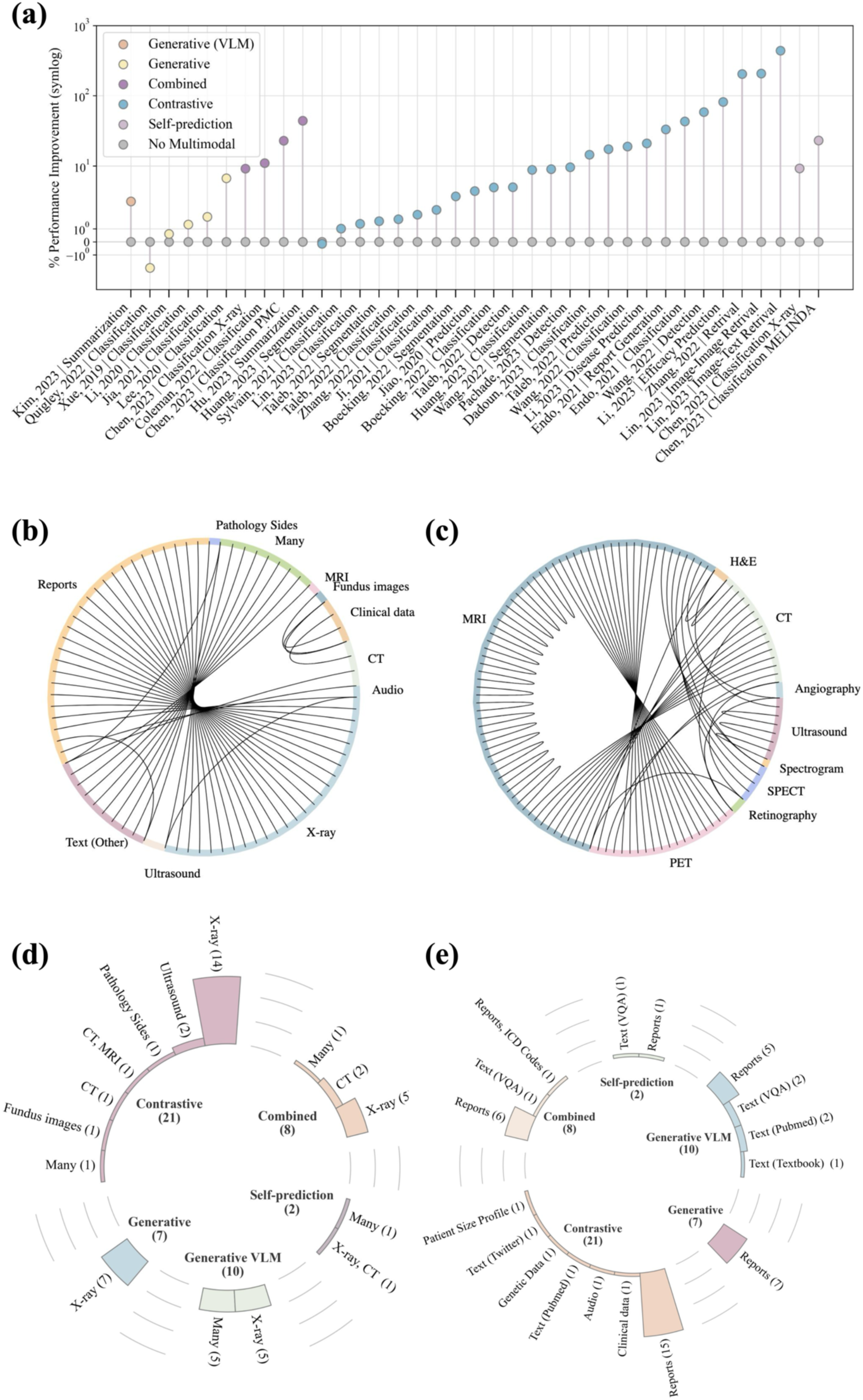
Summary of extracted data from studies in our systematic review. (a) Percentage improvement in downstream task performance using multimodal training compared to single modality approaches. (b) Pairing combinations of image and non-image data during pretraining. (c) Pairing combinations of different imaging modalities during pretraining. (d) Prevalence of imaging modalities across various pretraining types. (e) Prevalence of non-imaging modalities across various pretraining types.

**Table 1:**
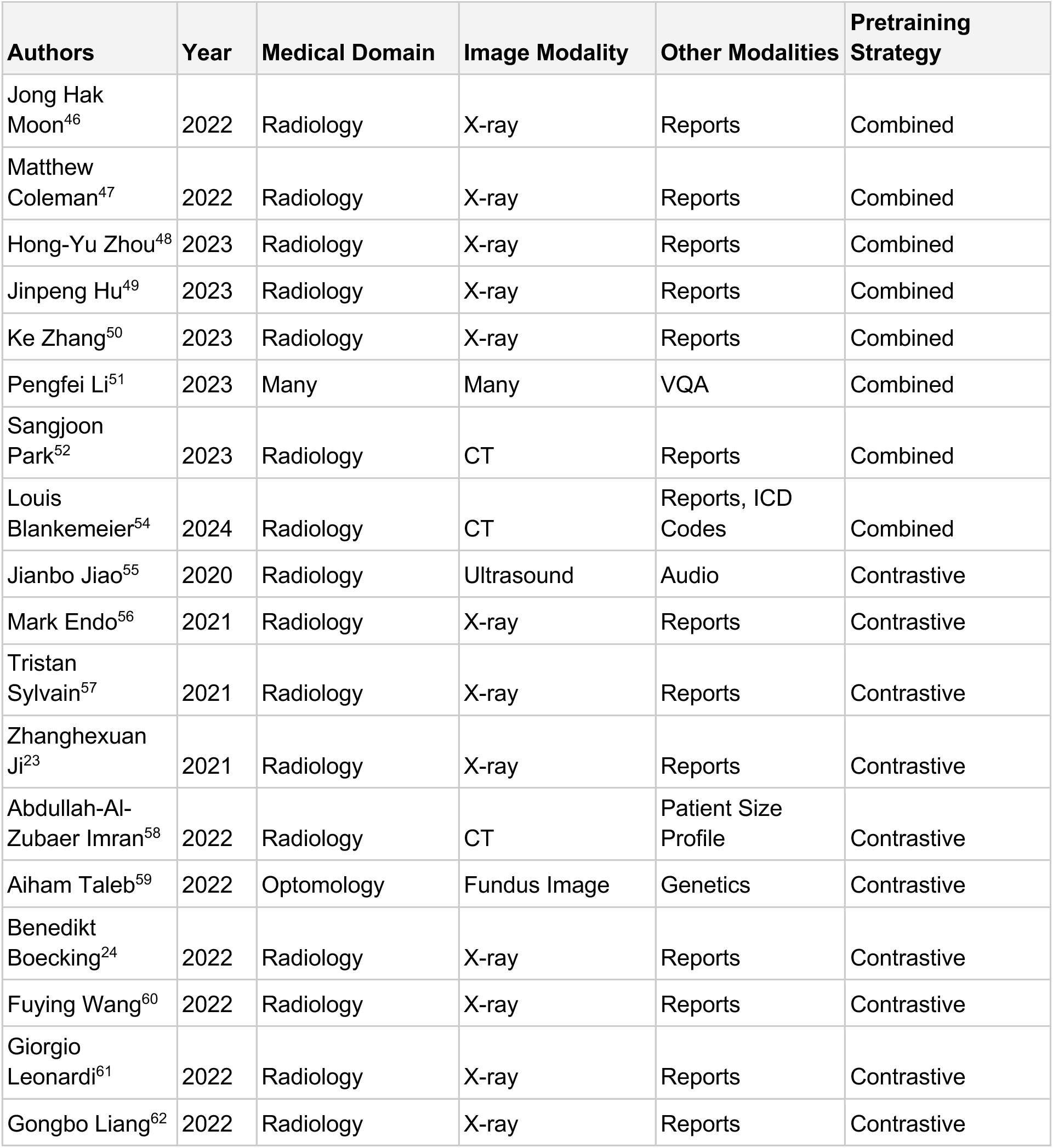
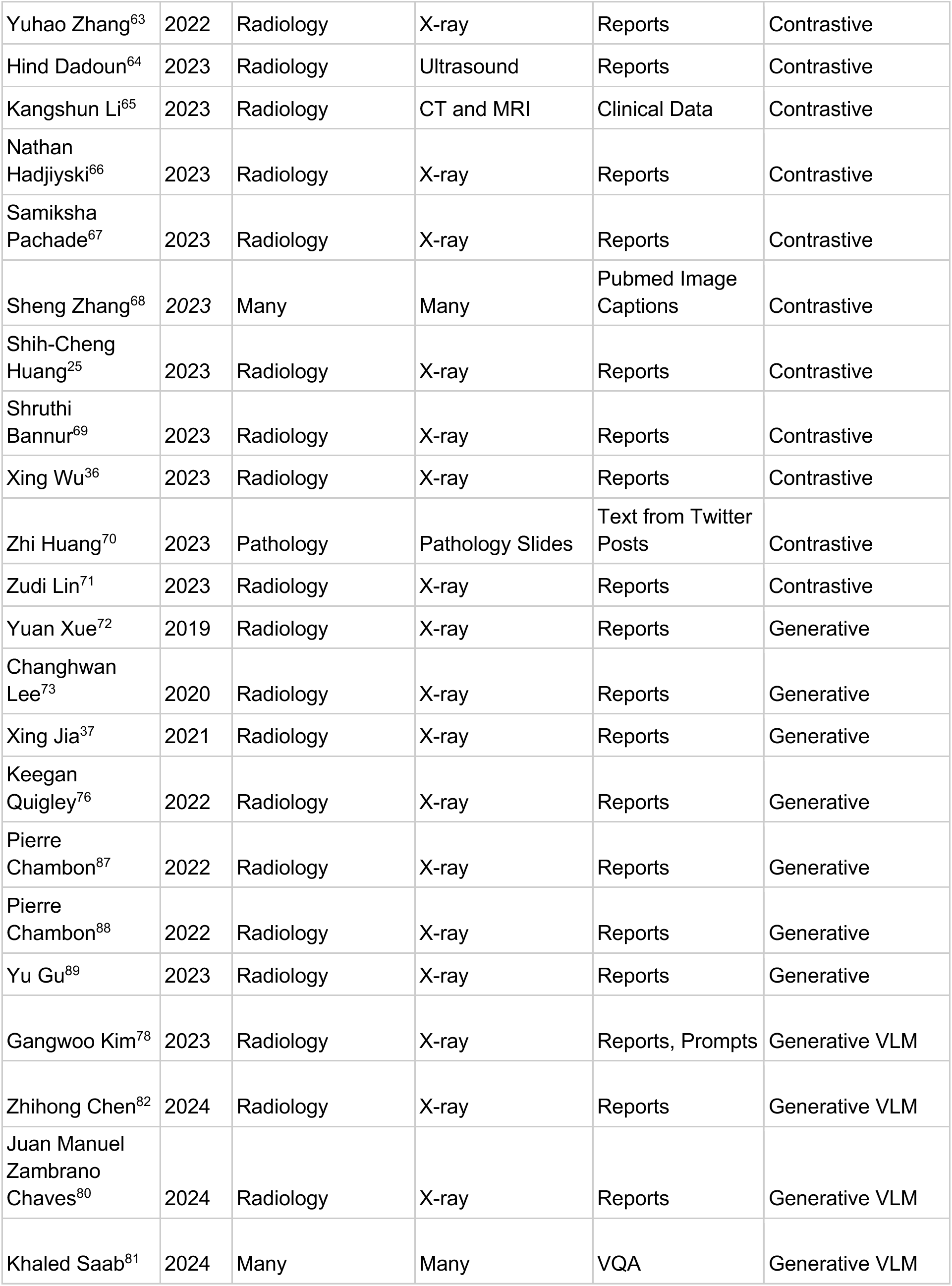
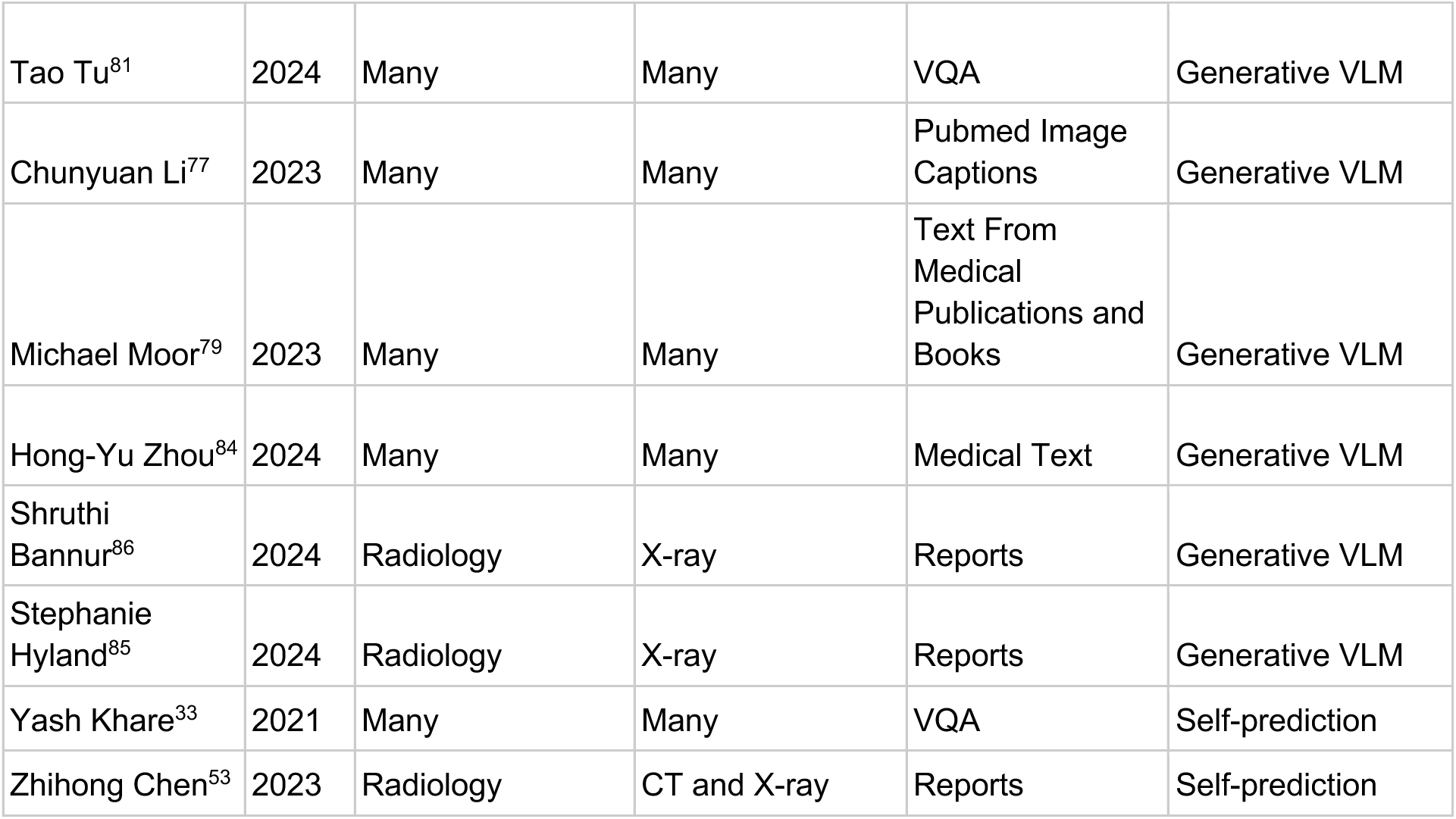
Overview of image to non-imaging multimodal self-supervised studies included in the systematic review.

**Table 2:** Overview of image to image multimodal studies included in the systematic review.

| Authors | Year | Medical Domain | Pretraining Strategy | Modalities |
| --- | --- | --- | --- | --- |
| Hong Liu<sup>92</sup> | 2023 | Radiology | Combined | MRI (T1, T1-C, T2, FLAIR) |
| Redha Touati<sup>93</sup> | 2023 | Radiology | Combined | MRI (T1, T2, T1ce, FLAIR) |
| Alex Fedorov<sup>94</sup> | 2021 | Radiology | Contrastive | MRI (T1, fALFF) |
| Ruobing Huang<sup>95</sup> | 2021 | Radiology | Contrastive | Ultrasound, B-mode, SWE, SE, Doppler |
| Yanbei Liu<sup>96</sup> | 2021 | Radiology | Contrastive | MRI, PET |
| Alex Fedorov<sup>97</sup> | 2023 | Radiology | Contrastive | MRI (T1, fALF) |
| Zheling Meng<sup>98</sup> | 2023 | Radiology | Contrastive | B-mode, Ultrasound elastography, Dynamic contrast-enhanced ultrasound |
| M. Jorge Cardoso<sup>99</sup> | 2015 | Radiology | Generative | MRI, CT |
| Yawen Huang<sup>100</sup> | 2016 | Radiology | Generative | MRI (T2, proton density) |
| Yawen Huang<sup>101</sup> | 2018 | Radiology | Generative | MRI (T1, T2, proton density) |
| Yongsheng Pan<sup>102</sup> | 2018 | Radiology | Generative | MRI, PET |
| Bo Peng<sup>103</sup> | 2019 | Radiology | Generative | MRI, Ultrasound |
| Wei wei<sup>104</sup> | 2019 | Radiology | Generative | MRI, PET |
| Aman Rana<sup>105</sup> | 2020 | Pathology | Generative | Deparaffinated slice, HE stained slice |
| Anmol Sharma<sup>106</sup> | 2020 | Radiology | Generative | MRI (T1, T2) |
| Haoyu Lan<sup>107</sup> | 2020 | Radiology | Generative | MRI, PET |
| Jaya Chandra Raju<sup>108</sup> | 2020 | Radiology | Generative | MRI (T2, T1(Gd), T2, Flair) |
| Jianbo Jiao<sup>109</sup> | 2020 | Radiology | Generative | Ultrasound, MRI |
| Jose Morano<sup>110</sup> | 2020 | Ophthalmology | Generative | Retinography, angiography |
| Xianjin Dai<sup>111</sup> | 2020 | Radiology | Generative | MRI (T1, T2, FLAIR) |
| Yongsheng Pan<sup>112</sup> | 2020 | Radiology | Generative | MRI, PET |
| Bo Zhan<sup>113</sup> | 2021 | Radiology | Generative | MRI (T1, T1c, T2, FLAIR) |
| Haohui Liu<sup>114</sup> | 2021 | Radiology | Generative | MRI (T1, FLAIR), PET |
| Lipei Zhang<sup>115</sup> | 2021 | Radiology | Generative | MRI, Low count PET, PET |
| Wanyun Lin<sup>116</sup> | 2021 | Radiology | Generative | MRI, PET |
| Yuchen Fei<sup>117</sup> | 2021 | Radiology | Generative | MRI (T1, T1c, T2, FLAIR) |
| Álvaro S. Hervella<sup>118</sup> | 2022 | Ophthalmology | Generative | Retinography, fluorescein angiography |
| Chao Fan<sup>119</sup> | 2022 | Radiology | Generative | MRI, CT, PET, SPECT |
| Hongfei Sun<sup>120</sup> | 2022 | Radiology | Generative | CBCT, CT, MRI |
| Jin Zhang<sup>121</sup> | 2022 | Radiology | Generative | MRI, PET |
| Jitao Li<sup>122</sup> | 2022 | Radiology | Generative | PET, CT, MRI |
| Lu Tang<sup>123</sup> | 2022 | Radiology | Generative | MRI, PET |
| Nahed Tawfik<sup>124</sup> | 2022 | Radiology | Generative | CT, MRI (T2), PET, MRA, SPECT |
| Onat Dalmaz<sup>125</sup> | 2022 | Radiology | Generative | MRI (T1, T2, FLAIR, proton density), CT |
| Shangwang Liu<sup>126</sup> | 2022 | Radiology | Generative | MRI, PET, SPECT |
| Sureerat Reaungamornrat<sup>127</sup> | 2022 | Radiology | Generative | CT, MRI |
| Suzhe Wang<sup>128</sup> | 2022 | Radiology | Generative | CT, perfusion CT |
| Wang A<sup>129</sup> | 2022 | Radiology | Generative | MRI, CT, PET |
| Xiofeng Liu<sup>130</sup> | 2022 | Radiology | Generative | Cine MRI, spectrogram |
| Bing Cao<sup>131</sup> | 2023 | Radiology | Generative | MRI (T1, T1-C, T2, FLAIR) |
| Jiaojiao Zhang<sup>132</sup> | 2023 | Radiology | Generative | CT, PET, MRI (T1CE, T2, FLAIR, T1) |
| Redha Touati<sup>133</sup> | 2023 | Radiology | Generative | MRI (T2, T1ce, FLAIR) |
| Weisheng Li<sup>134</sup> | 2023 | Radiology | Generative | MRI, CT, PET, SPECT |
| Xinyu Xie<sup>135</sup> | 2023 | Radiology | Generative | MRI, CT, PET, SPECT |
| Aiham Taleb<sup>136</sup> | 2021 | Radiology | Pretext Task | CT, MRI (T1, T1Gd, T2, FLAIR, ADC) |
| Zhuo Xiang<sup>137</sup> | 2022 | Radiology | Pretext Task | SWE, ultrasound, CDUS |
| Gijs van Tulder<sup>138</sup> | 2023 | Radiology | Pretext Task | MRI (T1, T1c, T2, FLAIR, Normal vs fat-suppressed) |
| Qingfan Hou<sup>139</sup> | 2023 | Radiology | Pretext Task | MRI (T1, T2, T1Gd, FLAIR) |
| Qian Zhou<sup>140</sup> | 2022 | Radiology | Self-prediction | MRI (T1, T2, FLAIR) |

### Contrastive

Contrastive SSL was utilized in 21 out of 48 studies (Table 1). For imaging modalities used in these studies, X-rays were the most prevalent, featured in 14 studies^23–25,36,56,57,60–63,66,67,69,71^. Ultrasound was used in 2 studies^55,64^, CT images in 1 study^58^, and a combination of CT and MRI in another^65^. Additionally, fundus images^59^, pathology slides^70^, and medical images from pubmed papers^68^ were each employed in 1 study. For the corresponding non-imaging modalities, radiology reports were the most common, appearing in 15 studies^23–25,36,56,57,60–64,66,67,69,71^, while 2 studies used other text sources, specifically PubMed image captions^68^ and text from Twitter posts^70^. Genetic data^59^, patient size profiles^58^, clinical data, and speech^55^ were each used once in separate studies.

Eight studies used traditional image-text contrastive learning similar to CLIP^36,56,62–64,67,68,70^, 7 studies employed global and local contrastive learning^23–25,57,60,61,66^, 1 study adopted a strategy akin to SimSiam^65^, 1 study explored local, global and temporal correspondence^69^ and 4 studies developed novel strategies^55,58,59,71^. The reported average improvement of multimodality over single modality, where available, was: 0.050 AUROC (10 studies^23–25,55,57,59,60,63,67,71^), 0.201 accuracy (1 study^65^), 0.362 Precision@5 (2 studies^63,71^), 0.023 BLEU 2 (1 study^56^), 0.103 F1-score (2 studies^56,64^), 0.028 Dice (3 studies^59,60,69^), and 0.092 mAP (1 study^60^)

While most studies apply contrastive learning between text and images, two studies applied contrastive learning to other combinations of modalities. Taleb et al. was the only study across all categories that combined images and genetic data^59^. They utilized fundus images in conjunction with Single Nucleotide Polymorphisms (SNP) and Polygenic Risk Scores from the UK Biobank to create positive pairs within each patient, using other patients as negative pairs. Their findings demonstrate that this method can enhance fundus pathology detection and facilitate the identification of genetic associations with fundus diseases. Jiao et al. utilized paired ultrasound images and audio of a clinician describing findings during the ultrasound^55^. They created positive pairs between speech and ultrasound at the same time points, while later time points served as negative pairs, and audio sections with background noise were used as hard negatives.

Two studies chose non-traditional approaches for collecting paired images and text^68,70^. Rather than using medical images and radiology reports, Zhang et. al. scraped PubMed for papers with medical images and their corresponding captions, yielding a dataset of over 15 million image-caption pairs^68^. The authors used CLIP style training to train BiomedCLIP, which outcompeted several benchmarks in VQA, image classification and retrieval. Similarly, instead of relying on publicly released datasets or proprietary hospital data, Huang et. al. used Twitter posts of pathology slices with their corresponding text to curate a public dataset^70^.

### Self-prediction

Two of the 48 studies utilized self-prediction as a pre-training method (Table 1). Khare et al. presented MMBERT^33^, which used masked language modeling to train a Visual Question Answering (VQA) model by masking out words and restoring the original caption, jointly utilizing both language and image features. They trained their model on the ROCO dataset, which contains various types of radiology images with corresponding question-answer pairs. They did not report single modality performance^33^. Chen et al. employed cross-attention between encoders in masked language modeling and masked image modeling to restore both text and images^53^. They demonstrated their methods on X-rays and CT scans, using radiology reports as their second modality. Their work reported an increase of 0.147 in accuracy and 0.075 in AUROC for multimodal over single-modality approaches.

### Generative

Generative approach was used in 7 out of 48 studies (Table 1). All generative papers used X-ray images as their imaging modality and radiology reports for their corresponding modality. Four out of the 7 papers pretrained their models based on radiology report findings generation^37,72,73,76^. The reported average improvement of multimodality over single modality was 0.002 AUROC (3 studies^37,72,76^) and 0.053 F1 score (1 study^73^). Notably, Quigley et al. reported a higher AUROC for their unimodal text-based model when using 100% of the training data for fine-tuning, but a higher AUROC for their multimodal approach when only a subset of the training data were used.

The remaining 3 pretrained their models by generating synthetic chest X-ray based on radiology reports or text prompts^87–89^. Chambon et al. demonstrated successful adaptation of a general domain image generation model, Stable Diffusion, to generate synthetic chest X-ray images^88^.

RoentGen further validated the utility of synthetic images by showing a 5% points improvement in classifier performance when trained jointly on synthetic and real images^87^. BiomedJourney showcased the capability to edit chest X-ray images using natural language instructions, enabling the creation of counterfactual images^89^. For instance, the model can generate specific abnormalities on a healthy patient’s chest X-ray using prompts, effectively transforming normal images into ones displaying requested pathologies.

### Generative VLM

Ten out of 48 studies^77–84^ leveraged existing text-based foundation models (LLMs) to develop VLMs using generative pretraining (Table 1). Out of the 10 studies, 5 studies specifically focused on X-ray as their imaging modality^78,80,82,85,86^, while the remaining 5 were capable of analyzing several different imaging modalities. In terms of the corresponding modality, 3 studies used the corresponding radiology reports^78,80,82,85,86^ while 2 uses questions from VQA datasets^81,83^. Three studies found creative ways to source corresponding text, including Pubmed image captions^77^ and text from publications and medical textbooks^79,84^. Of these studies, 1 reported an improvement of 1.45 ROUGE from multimodal training over single modality training^78^. The remaining studies did not perform this comparison.

Some notable work includes Med-PaLM Multimodal^83^, which emerged as a model capable of encoding and interpreting a wide array of biomedical data, including clinical language, imaging, and genomics, all with the same set of model weights. Med-Gemini^81^ further improves upon Med-PaLM’s capabilities by leveraging the capabilities of the large VLM Gemini^14^. In addition, Med-Gemini incorporated self-training and web search integration, enhancing the model’s ability to verify its outputs and improve reliability. MedVersa^84^ introduced an innovative approach using an orchestrator powered by a LLM to independently assess whether to execute tasks on its own or integrate visual modeling modules, potentially improving efficiency and accuracy in medical image analysis.

Several studies demonstrated that generative VLMs need not be proprietary; instead, smaller, open-source VLMs can rival the performance of their larger, closed-source counterparts^77,80,82,85,86,90^. LLaVA-Med^77^, demonstrated that the open-sourced model, LLaVA^13^, could be successfully adapted to the medical domain in less than a day of training using open-sourced model LLaVA^13^. LLaVA-Rad extends on LLaVA-Med to focus on the task of report generation, and further introduced a more clinically relevant metric, CheXprompt, which showed no statistically significant difference from human radiologist evaluations. Similar to LLaVA-Rad, MAIRA-2 is a small yet effective report generation model. Furthermore, MAIRA-2 demonstrated that generated reports can be grounded by associating generated text with bounding boxes, providing an easier way for physicians to verify the generated report based on visual signals in the image^86^.

### Combined

A combined SSL approach was employed in 8 out of the 48 studies (Table 1). Among these studies, 5 used X-rays as the imaging modality^46–50^, 2 used CT scans^52,54^, and 1 used multiple types of radiology images^51^. Regarding the non-imaging modality, 6 studies used radiology reports^46–50,52^, while 1 study used text from VQA^51^, and 1 used both report and ICD codes^54^. Contrastive learning emerged as the most frequently method in a combined pretraining strategy, being utilized in 6 out of the 8 studies that used a combined SSL approach^48–52,54^. In 3 of these 6 studies^50–52^, contrastive learning was combined with masked modeling, while in the remaining 2 studies^48,49^ it was combined with a generative task and 1 pretrained the model with a pretext task^91^ before contrastive learning^54^. One study employed masked language modeling and image-report matching together^46^, while another study utilized a diverse set of approaches^47^. Overall, an increase in AUROC of 0.098 (1 studies^47^) and 20.8 in ROUGE-L (1 study^49^) was reported for multimodality over single modality.

Notably, Sangjoon Park et al. introduced artificial errors in radiology reports and trained a model to detect these by combining CLIP-style contrastive learning, multi-modal masked modeling and momentum updating of a teacher model akin to DINO training^52^. Pengfei Li et al. 2023 combined contrastive learning, masked language modeling, and image text matching to pretrain on multiple large open medical image datasets, followed by fine tuning on VQA-RAD, PathVQA, SLAKE where their method exceed state-of-the-art on VQA^51^. Lastly, Blankemeier et al. used a creative approach of training a CT foundation model, Merlin, by first utilizing a pretext task^91^ of predicting ICD codes from CT scans and subsequently continue the training of the model with a CLIP-style contrastive objective between CTs and reports, allowing their model to achieve state-of-the-art performance on numerous tasks^54^. Notably, this study stands out as one of the few that focuses on ingesting and processing full CT scans, expanding the application of foundation models beyond the more commonly studied 2D modalities such as X-rays and addressing the unique challenges and opportunities presented by three-dimensional imaging data.

### Image-Image

Our review also identified 49 papers that employ multimodal self-supervised pretraining between two imaging modalities (Table 2). Among the imaging modalities used in these studies, MRI was _the_ most frequently employed (42/49)^92–94,96,97,99–104,106–109,111–117,119–127,129–136,138–140^), followed by PET (16/49 studies^96,102,104,107,112,116,119,121–124,126,129,132,134,135^), CT (13/49 studies)^99,119,120,122,124–129,132,134,135^, and ultrasound (5/49 studies^95,98,103,109,137^) Which modalities were combined in each study can be seen in Figure 5. The majority of these studies (37/49) adopted a generative approach to synthesize one imaging modality from another. One used self-prediction^140^, 5 used contrastive methods^94–98^, 2 used a combined approach^92,93^, and the remaining 4 used other approaches such as modality prediction^136–139^. In terms of medical specialty, neuroimaging is the most prominent area for image-image self-supervised pretraining, with 35 out of 49 studies focusing on this field. In the remaining studies^92,93,96,97,99–102,104,106–109,111–114,116,117,119,121–126,128,129,131–135,139,140^, 11 focused on radiology^94,95,98,103,115,120,127,130,136–138^, 2 on ophthalmology^110,118^, and 1 on pathology^105^. For many of these studies with downstream tasks, the main applications are Alzheimer’s disease diagnosis (5 studies^94,96,97,102,116^) and brain tumor segmentation (5 studies^92,93,132,136,139^).

## Discussion

The purpose of this systematic review is to synthesize the current state of knowledge on the application of multimodal foundation models for medical imaging. We propose a unified terminology for multimodal self-supervised methods and a taxonomy for prior work based on their pretraining strategies. We screened 1,144 papers from medical and AI domains and extracted data from 97 included studies. Based on our review, we found that multimodal self-supervised pretraining generally improves downstream task performance compared to single modality pretraining, with gains ranging up to 439% across studies (Figure. 5). The nascent nature of the multi-modal deep learning field and the heterogeneity in experimental setups currently precludes definitive conclusions about the superiority of specific multimodal self-supervised learning strategies across all medical imaging domains and modalities. Despite these limitations, our findings suggest that multimodal self-supervised pretraining is a promising approach for enhancing medical image models, and we encourage researchers to explore these techniques in future work.

Our systematic review reveals an emerging trend towards generative VLMs that leverage the advanced capabilities of LLMs for medical tasks. These large models, typically many billions of parameters, demonstrate extensive versatility in handling diverse modalities and performing a wide array of downstream tasks. For instance, Med-PaLM multimodal^83^ showcases the ability to process and interpret biomedical data spanning clinical language, imaging, and genomics, all within a unified model architecture. The trend towards these comprehensive generative models is not only driven by their performance and generalizability but also by their natural language interactive interfaces. These chat-like features enable nuanced physician-AI collaboration and discussion for diagnosis, moving beyond simple reliance on AI outputs to potentially improve patient outcomes.

It is crucial to acknowledge that the sheer scale and computational demands of these models can preclude their deployment within hospital firewalls, raising legitimate concerns about the privacy and security of patient health records when transmitted to external model providers. Addressing this challenge, foundation models such as LLaVA-Rad^80^ and CheXagent^82^, which are smaller and open-source, rival the capabilities of their larger counterparts, making local deployment more feasible. Furthermore, advances in foundation model development must be coupled with some challenges such as their tendency to hallucinate, which could lead to potentially harmful misdiagnoses in healthcare settings. The approach taken by models like Med-Gemini^81^, which incorporates online search capabilities to verify its outputs, sets a valuable precedent for enhancing the reliability and safety of AI-assisted medical decision-making. These considerations underscore the importance of balancing model capability, deployability, and safety as we continue to develop and refine foundation models for healthcare applications.

Training these foundation models typically requires vast amounts of unlabeled data. This substantial data volume and diverse dataset not only contributes to the model’s emerging properties but has also been shown to improve the model’s resilience to distribution shift^141^. Such robustness is crucial for deployment in hospital settings, where variations in imaging equipment or patient populations can lead to significant distributional changes. However, patient privacy concerns often restrict access to large-scale medical data, creating a significant hurdle in the development process. The largest publicly accessible medical datasets^142–145^ pale in comparison to the internet-scale data used to train general domain foundation models. In response to this challenge, several studies in our review have identified innovative approaches to data sourcing, such as leveraging PubMed images and captions^68^, extracting interleaved text and images from medical textbooks^79^, and even mining relevant posts from social media platforms like Twitter^70^. As we move forward in developing more powerful and generalizable medical AI models, these innovative data collection and pairing techniques will likely play an increasingly crucial role in overcoming limitations posed by data scarcity and privacy concerns. However, developers must also be aware that while large-scale datasets from public sources can provide valuable training data, they may not always meet the rigorous standards required for clinical applications. Therefore, future model development must carefully navigate the tradeoff between data quantity and quality, balancing the benefits of large-scale datasets with the need for high-quality, clinically relevant information to ensure both powerful and reliable AI models for real-world medical applications.

While many of the papers in our review focused on developing multimodal foundation models using medical images and text, it is crucial to emphasize that these models should expand beyond natural languages to better align with the multifaceted nature of healthcare. Our review identified several studies that incorporated unique modalities during self-supervised pretraining, including genetic data^59^, clinical data^65^, ICD codes^54^, speech^55^, and patient size profiles^58^. The inclusion of these diverse modalities provides the model with a more comprehensive view of the patient, mirroring the approach taken by physicians in clinical practice. This multi-modal approach not only enhances the model’s diagnostic and prognostic capabilities but also opens new avenues for discovering complex associations between different modalities. For instance, the ability to correlate genetic data with pathology slides could uncover intricate relationships that might be challenging for human experts to identify independently. Such capabilities have the potential to significantly create new opportunities for research in fields such as biology and treatment development. As the field of medical AI continues to advance, it is imperative to develop truly comprehensive multimodal models that can integrate and analyze the full spectrum of patient data available in modern healthcare settings.

Our systematic review also identified 49 studies that utilize multimodal self-supervised pretraining between two imaging modalities (Table 2). While these methods demonstrate utility in specific medical applications, such as image registration, denoising, and reconstruction, it is important to note their limitations. Notably, less than half of these studies (23/49) rigorously evaluate the effectiveness of their learned multimodal representations on downstream tasks, which raises questions about their generalizability and label efficiency. More critically, the focus on image-to-image modalities alone may be limiting the potential of these models. As mentioned above, recent advancements in vision foundation models have demonstrated that their capabilities are significantly enhanced when paired with text or other non-imaging modalities. This suggests that while image-to-image multimodal learning has its place in specific medical imaging tasks, the future of medical AI likely lies in more comprehensive multimodal approaches that incorporate diverse data types.

### Guidelines

Our systematic review demonstrates the significant potential of multimodal self-supervised pretraining in enhancing medical imaging AI models across various domains, modalities, and downstream tasks. However, we also observe that the clinical adoption of these advanced models remains limited. Several factors contribute to this gap between technological capability and practical implementation, including potential biases in model predictions, insufficient demonstration of clinical utility, and inadequate consideration of implementation challenges. To bridge this divide and facilitate the responsible integration of these powerful AI tools into healthcare, a collaborative effort among all stakeholders is crucial. Model developers, clinicians, policymakers, and dataset curators must work in concert to address the multifaceted challenges identified in our review. Recognizing the critical importance of this interdisciplinary collaboration, we provide targeted guidelines for each of these key players below. These recommendations aim to foster a cohesive approach to developing AI systems that are not only technically sophisticated but also clinically relevant, ethically sound, and practically implementable in real-world healthcare settings.

#### Guidelines for Model Developers

Model developers should leverage the recent advances in multimodal self-supervised learning techniques from the general domain when building medical imaging AI models. However, it is crucial to consider the unique properties and differences between general domain images and medical images when applying these methods^91^. One key difference is that, unlike natural images where class-defining features often occupy a significant portion of the image, medical images typically have more localized and subtle class-defining features, such as abnormalities. Consequently, popular multimodal self-supervised methods like CLIP^22^, which rely on learning joint representations between global image and text features, may have limitations in capturing these subtle, localized features in medical images. To address this challenge, innovative approaches have been proposed to adapt methods from the general domain to the specific characteristics of medical images. For example, GLoRIA^25^, ViLLA^146^, and BioViL^147^ demonstrate a promising approach to modify self-supervised learning techniques to better suit the unique properties of medical imaging data. Future developers should also consider introducing technical innovations to adapt general domain methods to meet the specific challenges and characteristics of medical images.

In addition to developing medical imaging-specific methods, developers should also consider using evaluation metrics tailored to specific medical tasks. For instance, in radiology report generation, two common types of metrics are used: (1) lexical similarity-based metrics (i.e. BLUE^148^, ROUGE^149^), which assess whether the model’s outputs are contextually and stylistically aligned with human-written reports, and (2) factual correctness metrics (i.e. F1-CheXpert^143^, F1-RadGraph^150,151^), which evaluate the extent to which the generated reports accurately reflect the imaging findings. While both coherence and factual accuracy are essential for high-quality radiology reports, studies have found that these metrics have limited correlation with manual error scoring performed by radiologists, which is more clinically useful^152^. To address this discrepancy, researchers have proposed novel approaches to automatically evaluate radiology report generation models. For example, the LLaVA-Rad^80^ introduced a method that uses GPT-4 to analyze the error types in the generated reports automatically. The resulting metric, CheXpromt, has been shown to have no statistically significant difference compared to human radiologist evaluations, suggesting that it could be used as a substitute for manual radiologist assessment when evaluating the clinical utility of these models. Furthermore, methods like GREEN^153^ have shown that GPT-4’s knowledge can be distilled into smaller, open-source models for report evaluation, eliminating the need for API calls and enhancing accessibility and efficiency for researchers and developers. Moving forward, future work should prioritize the development and adoption of metrics that are more closely aligned with clinical relevance and utility.

#### Guidelines for Clinicians

Building clinically useful medical AI models is not solely the responsibility of model developers; clinicians play a crucial role and should actively collaborate with AI teams^4^. Often, models are developed based on the availability of datasets rather than addressing a genuine clinical need. Consequently, even if these models achieve high evaluation metrics, their utility may be limited in the absence of a clear clinical application. To ensure the development of clinically relevant AI models, it is essential for clinicians to identify true needs in healthcare settings that can be fulfilled or enhanced by AI. Once a clinical need is identified, clinicians should also identify the modalities that are required to complete the task. Lastly, physicians should determine a specific “action” to pair with the machine learning model’s output to address this need effectively. By defining a “decision-action” pair^154^, AI developers can evaluate the model’s utility based on the estimated net benefit in the context of the clinical need. Identifying a genuine clinical need, the modalities required to address this need, and defining an appropriate decision-action pair are instrumental in creating useful and deployable medical AI models, underscoring the importance of clinician involvement throughout the entire AI model development and deployment process.

Once AI models are deployed in clinical settings, it is imperative for clinicians and healthcare providers to maintain a critical and vigilant approach to their utilization. While these foundation models demonstrate impressive capabilities and are continuously improving, they are not yet sufficiently advanced to operate autonomously in healthcare environments. Clinicians should remain acutely aware of the models’ limitations, including their propensity for hallucination and susceptibility to performance degradation due to distribution shifts. It is crucial for healthcare professionals to actively monitor the models’ outputs, identifying and documenting any errors or inconsistencies observed during clinical use. Establishing a robust feedback loop between clinicians and model developers is essential for the continuous improvement and refinement of these AI systems. In this context, the emergence of VLMs with interactive chat interfaces presents a valuable opportunity for more nuanced clinician-AI collaboration in diagnostic processes. These interfaces enable healthcare providers to engage in detailed discussions with the AI, potentially uncovering insights or limitations that might not be apparent in more rigid, output-only systems. By fostering this type of interactive and critical engagement with AI tools, clinicians can play a pivotal role in enhancing the reliability, safety, and effectiveness of AI in healthcare, ultimately ensuring that these advanced technologies serve as powerful adjuncts to, rather than replacements for, human clinical expertise.

#### Guidelines for Policy Makers

Policy makers play a crucial role in shaping the development and deployment of medical imaging AI, particularly in the context of multimodal foundation models. To enable responsible innovation while ensuring patient safety, policy interventions should focus on several key areas. First, policy makers should consider establishing an expedited approval pathway for approved multimodal foundation models adapting to unapproved clinical tasks, similar to the FDA’s 510(k) process, to facilitate efficient deployment while maintaining stringent safety standards. This approach is warranted by the demonstrated capacity of foundation models to generalize to novel tasks with minimal additional training data. The approval process should distinguish between models utilizing previously approved modalities for inference and those incorporating entirely new modalities, with the latter necessitating more comprehensive evaluation. Secondly, policies should mandate sensitivity analyses on combinations of input modalities to ensure developers thoroughly assess model performance across various modality combinations and sources, addressing potential performance variability. This requirement is critical as model behavior may fluctuate based on available input modalities, and not all modalities may be present in real-world clinical scenarios. For example, a newly admitted patient might lack sufficient clinical history or a specific medical imaging modality that a particular foundation model was trained on. Understanding model behavior in these cases is crucial to prevent unexpected or potentially harmful predictions. Lastly, as generative AI models become increasingly prevalent in medical imaging applications, policy makers should develop comprehensive evaluation guidelines for tasks generative tasks, such as clinical report generation or summarization. As mentioned in the “Guidelines for Model Developers” section, traditional lexical similarity or factual correctness NLP metrics may be inadequate for evaluating generated medical text. This could involve establishing standards for human expert evaluation of generated reports or incorporating AI-assisted judgment systems, as demonstrated to be feasible in studies like LLaVA-Rad (Chaves et al., 2024). A two-tiered evaluation system, involving both human physicians and AI, with a mechanism for resolving discrepancies, could enhance the reliability, clinical relevance and feasibility of these assessments. By addressing these areas, policy makers can foster an environment that promotes the responsible development and implementation of advanced AI models in medical imaging, ultimately leading to improved patient care and outcomes.

#### Guidelines for Dataset Curators

Many existing publicly available medical image datasets are sourced primarily from developed countries, which can lead to biases that disproportionately affect model performance when deployed in developing countries or among minority groups in developed regions, where data representation may be inadequate^155–157^. To mitigate these biases, dataset curators need to prioritize the collection and inclusion of data from diverse populations. They should make a concerted effort to curate data from a wide range of demographics, ensuring that the dataset is representative of the global population. Furthermore, dataset curators should include patient demographic information in the dataset to enable model developers to evaluate the fairness and generalizability of their developed models across different subgroups. By taking these steps to curate diverse and inclusive datasets, we can help ensure that medical AI models are trained and evaluated on representative data and can provide equitable benefits to patients across different regions and demographics. This approach will help ensure that the benefits of AI-driven healthcare are distributed equitably across different populations and regions.

## Conclusions

In conclusion, our systematic review highlights the significant potential of multimodal foundation models in advancing medical imaging and healthcare AI. These models, particularly those leveraging vision and language capabilities, demonstrate promising improvements in performance and generalizability across various medical tasks. However, their development and deployment face challenges, including data scarcity, privacy concerns, and the need for interpretability and safety in clinical settings. Moving forward, we advocate for research that focuses on: (1) developing smaller, privacy-preserving, and deployable models without compromising performance; (2) innovative data collection strategies that respect patient privacy; (3) incorporation of diverse non-imaging modalities to better reflect the complexity of healthcare; and (4) rigorous evaluation of these models on clinically relevant downstream tasks. As the field progresses, we anticipate that multimodal foundation models will play an increasingly crucial role in healthcare, potentially revolutionizing diagnosis, treatment planning, and patient care. However, their successful integration into clinical practice will require continued collaboration between AI researchers, healthcare professionals, and policymakers to ensure these powerful tools are developed and used responsibly, effectively, and ethically.

## Limitations

A notable constraint arises from the inherent publication bias within the extant literature, which predominantly features studies reporting positive outcomes. Such bias may inadvertently lead to an inflated perception of the efficacy associated with multi-modal self-supervised learning techniques. Our examination was deliberately confined to literature published subsequent to the year 2012, thereby excluding works that predate the advent of deep learning in the realm of computer vision. The heterogeneity presented in the methodologies of the reviewed studies, encompassing diverse imaging modalities, varied performance metrics, and distinct research objectives, precludes a comprehensive quantitative synthesis or direct comparison of the relative benefits conferred by different learning strategies. Moreover, the classification of multimodal self-supervised learning approaches within each analyzed study was subject to a certain level of subjectivity, especially in instances involving innovative, non-traditional, or hybrid methodologies. Furthermore, the selection criteria for studies were specifically tailored to the domain of medical images. This focus inherently limits the breadth of our review, overlooking the versatility of self-supervised pretrained models, which hold significant promise across a spectrum of other modalities. This delineation of limitations underscores the nuanced challenges encountered in synthesizing the current landscape of multi-modal self-supervised learning within medical imaging, and it delineates a clear avenue for future research endeavors.

## Data Availibility

The authors affirm that all data underpinning the conclusions of this research are accessible within the manuscript and its Supplementary Information.

## Methods

This systematic review was conducted based on the PRISMA guidelines^158^.

### Search Strategy

A systematic literature search was conducted in the two literature databases: PubMed and Scopus. For potentially eligible studies cited by articles already included in this review, additional targeted free-text searches were conducted on Google Scholar if they did not appear in Scopus or PubMed. The key search terms were based on a combination of three major themes: “self-supervised learning”, “medical imaging modalities” and “other medical modalities / multimodal”. Search terms for medical imaging were not limited to radiological imaging but were also broadly defined to include imaging from all medical fields, i.e., fundus photography, whole slide imaging, endoscopy, echocardiography. The search encompassed papers published between January 2012 and January 2024. The start date was considered appropriate due to the rising popularity of deep learning for computer vision since the 2012 ImageNet challenge. The complete search string for all three databases is provided in Supplementary Methods.

We **included** all research papers in English that used multi-modal self-supervision techniques to develop models for medical imaging tasks. We **excluded** studies that used non-human medical imaging data (i.e., veterinarian medical images). Studies that rely on derived imaging characteristics, including biomarkers and radiomic features, as opposed to utilizing raw images directly, are also excluded from consideration. Conference abstracts, review articles, letters to the editor, and any submissions not constituting original research were also excluded. Additionally, studies not centered on medical imaging, not employing self-supervision techniques, or not incorporating multimodal approaches were not considered. Papers focusing solely on image registration were also outside the scope of this review.

For papers that leverage medical images with another non-imaging modality (Table 1), we further constrained our inclusion criteria to studies that applied the self-supervised pretrained models to a downstream medical image task. In other words, it was not sufficient for the study to have merely developed a multimodal self-supervised pretrained model; the model had to be evaluated on a clinically relevant task using medical images. We defined a clinically relevant task as one that directly relates to a clinical application or has the potential to inform clinical decision-making. For example, the downstream task of classifying the frame number in a temporal sequence of frames from echocardiography was not considered a clinically relevant task, as it does not provide meaningful information to a clinician or patient that can be used in patient care.

### Study Selection

The Covidence software (www.covidence.org) was used for screening and study selection. After removing duplicates, studies were screened based on title and abstract. Subsequently, full texts were obtained and assessed for inclusion and data extraction. Study selection was performed by two independent researchers (S.-C.H., M.E.K.J.), and disagreements were resolved through discussion. In cases where consensus could not be achieved a third arbitrating researcher was consulted (A.S.C.).

### Data extraction

For benchmarking the existing **imaging & non-imaging approaches (Table 1**) we extracted the following data from each of the selected articles: (a) first author, (b) year of publication, (c) medical domain, (d) imaging modalities, (e) other non-image modalities, (f) pretraining strategy. We classified the specific multi-model self-supervised learning strategy based on the definitions in the “Terminology and Strategies” section.

For benchmarking the existing **imaging & imaging approaches (Table 2**) we extracted the following data from each of the selected articles: (a) first author, (b) year of publication, (c) medical domain, (e) pretraining strategy, (f) input modalities. We classified the pretraining strategies for these studies based on the definitions in the “Terminology and Strategies” section.

We extracted AUROC whenever this metric was reported, otherwise, we prioritized F1 score over accuracy and sensitivity. For NLP tasks, we prioritize longer subsequences (i.e. ROUGE-2 over ROUGE-11, ROUGE-L over ROUGE-2, etc.). We used ROUGE over BLEU due to its recall-oriented nature, which is crucial for capturing all relevant medical information. When the article contained results from multiple models (i.e., ResNet and Vision Transformer) on the same task, metrics from the experiment with the best-performing model were extracted. When the authors presented results on multiple clinical tasks, we extracted metrics for each of the downstream tasks. In instances where a particular clinical task was evaluated across several datasets, we selected the highest performance from among the datasets. Single-modality baseline performance, model architecture, pre-training dataset, and initialization were extracted when available in the experiments.

We provide in **Supplementary Table 1** all data extracted for imaging & non-imaging approaches, including full paper title, pretraining dataset, dataset size, image encoder, other modalities encoder, imaging model weight initialization, and other modalities model weight initialization. We provide in **Supplementary Table 2** all data extracted for imaging & imaging approaches including full paper title and downstream task.

## Data Availability

https://docs.google.com/spreadsheets/d/1KAuikeKjxh1O7z6dfdxRB8Mg1b2Q1oijSN8oyAFjFoE/edit?usp=sharing

https://docs.google.com/spreadsheets/d/1ZSPxzMIO-YhiilKsFgW2ZeArgd1GI_1ix5K8lE_ygsw/edit?gid=0#gid=0

## Bibliography

1. Roberts, M. et al. Common pitfalls and recommendations for using machine learning to detect and prognosticate for COVID-19 using chest radiographs and CT scans. Nature Machine Intelligence 3, 199–217 (2021).

2. Wynants, L. et al. Prediction models for diagnosis and prognosis of covid-19: systematic review and critical appraisal. BMJ 369, m1328 (2020).

3. Yang, Y., Zhang, H., Gichoya, J. W., Katabi, D. & Ghassemi, M. The limits of fair medical imaging AI in real-world generalization. Nat. Med. (2024) doi:10.1038/s41591-024-03113-4.

4. Huang, S.-C. et al. Developing medical imaging AI for emerging infectious diseases. Nat. Commun. 13, 7060 (2022).

5. Huang, S.-C., Pareek, A., Seyyedi, S., Banerjee, I. & Lungren, M. P. Fusion of medical imaging and electronic health records using deep learning: a systematic review and implementation guidelines. NPJ Digit Med 3, 136 (2020).

6. Cohen, M. D. Accuracy of information on imaging requisitions: does it matter? J. Am. Coll. Radiol. 4, 617–621 (2007).

7. Boonn, W. W. & Langlotz, C. P. Radiologist use of and perceived need for patient data access. J. Digit. Imaging 22, 357–362 (2009).

8. LeCun, Y. & Misra, I. Self-supervised Learning: The Dark Matter of Intelligence. Preprint at (2021).

9. Bommasani, R., et al. On the Opportunities and Risks of Foundation Models. arXiv [cs.LG] (2021).

10. OpenAI et al. GPT-4 Technical Report. arXiv [cs.CL] (2023).

11. Dubey, A., et al. The Llama 3 Herd of Models. arXiv [cs.AI] (2024).

12. Yang, Z., et al. The Dawn of LMMs: Preliminary Explorations with GPT-4V(ision). ArXiv abs/2309.17421, (2023).

13. Liu, H., Li, C., Wu, Q. & Lee, Y. J. Visual Instruction Tuning. arXiv [cs.CV] (2023).

14. Gemini Team et al. Gemini: A Family of Highly Capable Multimodal Models. arXiv [cs.CL] (2023).

15. Krishnan, R., Rajpurkar, P. & Topol, E. J. Self-supervised learning in medicine and healthcare. Nat Biomed Eng 6, 1346–1352 (2022).

16. Moor, M. et al. Foundation models for generalist medical artificial intelligence. Nature 616, 259–265 (2023).

17. Meskó, B. & Görög, M. A short guide for medical professionals in the era of artificial intelligence. NPJ Digit Med 3, 126 (2020).

18. Khan, W., et al. A Comprehensive Survey of Foundation Models in Medicine. arXiv [cs.LG] (2024).

19. Moher, D. et al. Preferred reporting items for systematic review and meta-analysis protocols (PRISMA-P) 2015 statement. Syst. Rev. 4, 1 (2015).

20. Chen, T., Kornblith, S., Norouzi, M. & Hinton, G. A Simple Framework for Contrastive Learning of Visual Representations. arXiv [cs.LG] (2020).

21. He, K., Fan, H., Wu, Y., Xie, S. & Girshick, R. Momentum Contrast for Unsupervised Visual Representation Learning. arXiv [cs.CV] (2019).

22. Radford, A., et al. Learning Transferable Visual Models From Natural Language Supervision. arXiv [cs.CV] (2021).

23. Ji, Z. et al. Improving Joint Learning of Chest X-Ray and Radiology Report by Word Region Alignment. Mach Learn Med Imaging 12966, 110–119 (2021).

24. Boecking, B. et al. Making the Most of Text Semantics to Improve Biomedical Vision– Language Processing. in Computer Vision – ECCV 2022 1–21 (Springer Nature Switzerland, 2022).

25. Huang, S.-C., Shen, L., Lungren, M. P. & Yeung, S. GLoRIA: A multimodal global-local representation learning framework for label-efficient medical image recognition. in 2021 IEEE/CVF International Conference on Computer Vision (ICCV) (IEEE, 2021). doi:10.1109/iccv48922.2021.00391.

26. Devlin, J., Chang, M.-W., Lee, K. & Toutanova, K. BERT: Pre-training of Deep Bidirectional Transformers for Language Understanding. arXiv [cs.CL] (2018).

27. Pathak, D., Krahenbuhl, P., Donahue, J., Darrell, T. & Efros, A. A. Context Encoders: Feature Learning by Inpainting. arXiv [cs.CV] (2016).

28. Dominic, J., et al. Improving Data-Efficiency and Robustness of Medical Imaging Segmentation Using Inpainting-Based Self-Supervised Learning. Bioengineering (Basel) 10, (2023).

29. Dosovitskiy, A., et al. An Image is Worth 16×16 Words: Transformers for Image Recognition at Scale. arXiv [cs.CV] (2020).

30. Bao, H., Dong, L., Piao, S. & Wei, F. BEiT: BERT Pre-Training of Image Transformers. arXiv [cs.CV] (2021).

31. He, K., et al. Masked Autoencoders Are Scalable Vision Learners. arXiv [cs.CV] (2021).

32. Li, Y., Fan, H., Hu, R., Feichtenhofer, C. & He, K. Scaling language-Image Pre-training via masking. Proc. IEEE Comput. Soc. Conf. Comput. Vis. Pattern Recognit. 23390–23400 (2022).

33. Khare, Y. et al. MMBERT: Multimodal BERT Pretraining for Improved Medical VQA. Proc. IEEE Int. Symp. Biomed. Imaging 1033–1036 (2021).

34. Bank, D., Koenigstein, N. & Giryes, R. Autoencoders. arXiv [cs.LG] (2020).

35. Suzuki, M. & Matsuo, Y. A survey of multimodal deep generative models. arXiv [cs.LG] (2022).

36. Wu, X., Li, J., Wang, J. & Qian, Q. Multimodal contrastive learning for radiology report generation. J. Ambient Intell. Humaniz. Comput. 14, 11185–11194 (2023).

37. Jia, X. et al. Radiology report generation for rare diseases via few-shot Transformer. Bioinform Biomed 1347–1352 (2021).

38. Goodfellow, I. J. et al. Generative Adversarial Networks. arXiv [stat.ML] (2014).

39. Ho, J., Jain, A. & Abbeel, P. Denoising Diffusion Probabilistic Models. arXiv [cs.LG] (2020).

40. Li, C., et al. Multimodal Foundation Models: From Specialists to General-Purpose Assistants. arXiv [cs.CV] (2023).

41. BLIP: PyTorch Code for BLIP: Bootstrapping Language-Image Pre-Training for Unified Vision-Language Understanding and Generation. (Github).

42. Yu, J., et al. CoCa: Contrastive Captioners are Image-Text Foundation Models. arXiv [cs.CV] (2022).

43. Zhang, S., et al. Instruction Tuning for Large Language Models: A Survey. arXiv [cs.CL] (2023).

44. Wei, J., et al. Finetuned Language Models Are Zero-Shot Learners. arXiv [cs.CL] (2021).

45. Kojima, T., Gu, S. S., Reid, M., Matsuo, Y. & Iwasawa, Y. Large language models are zero-shot reasoners. Adv. Neural Inf. Process. Syst. 35, 22199–22213 (2022).

46. Moon, J. H., Lee, H., Shin, W. & Choi, E. Multi-modal Understanding and Generation for Medical Images and Text via Vision-Language Pre-Training. arXiv [cs.CV] (2021).

47. Coleman, M., Dipnall, J. F., Jung, M. C. & Du, L. PreRadE: Pretraining Tasks on Radiology Images and Reports Evaluation Framework. Sci. China Ser. A Math. 10, 4661 (2022).

48. Zhou, H.-Y., et al. Generalized Radiograph Representation Learning via Cross-supervision between Images and Free-text Radiology Reports. arXiv [eess.IV] (2021).

49. Hu, J., Chen, Z., Liu, Y., Wan, X. & Chang, T.-H. Improving Radiology Summarization with Radiograph and Anatomy Prompts. arXiv [cs.CV] (2022).

50. Zhang, K. et al. Multi-Task Paired Masking With Alignment Modeling for Medical Vision-Language Pre-Training. IEEE Trans. Multimedia 26, 4706–4721 (2024).

51. Li, P., Liu, G., He, J., Zhao, Z. & Zhong, S. Masked Vision and Language Pre-training with Unimodal and Multimodal Contrastive Losses for Medical Visual Question Answering. in Medical Image Computing and Computer Assisted Intervention – MICCAI 2023 374–383 (Springer Nature Switzerland, 2023).

52. Park, S., Lee, E. S., Shin, K. S., Lee, J. E. & Ye, J. C. Self-supervised multi-modal training from uncurated images and reports enables monitoring AI in radiology. Med. Image Anal. 91, 103021 (2024).

53. Chen, Z. et al. Mapping medical image-text to a joint space via masked modeling. Med. Image Anal. 91, 103018 (2024).

54. Blankemeier, L., et al. Merlin: A Vision Language Foundation Model for 3D Computed Tomography. arXiv [cs.CV] (2024).

55. Jiao, J. et al. Self-Supervised Contrastive Video-Speech Representation Learning for Ultrasound. in Medical Image Computing and Computer Assisted Intervention – MICCAI 2020 534–543 (Springer International Publishing, 2020).

56. Endo, M., Krishnan, R., Krishna, V., Ng, A. Y. & Rajpurkar, P. Retrieval-Based Chest X-Ray Report Generation Using a Pre-trained Contrastive Language-Image Model. in Proceedings of Machine Learning for Health (eds. Roy, S. et al.) vol. 158 209–219 (PMLR, 2021).

57. Sylvain, T. et al. CMIM: Cross-modal information maximization for medical imaging. Proc. IEEE Int. Conf. Acoust. Speech Signal Process. 1190–1194 (2021).

58. Imran, A.-A.-Z. et al. Multimodal Contrastive Learning for Prospective Personalized Estimation of CT Organ Dose. in Medical Image Computing and Computer Assisted Intervention – MICCAI 2022 634–643 (Springer Nature Switzerland, 2022).

59. Taleb, Kirchler & Monti. ContIG: Self-supervised Multimodal Contrastive Learning for Medical Imaging with Genetics. Proc. IEEE.

60. Wang, F., Zhou, Y., Wang, S., Vardhanabhuti, V. & Yu, L. Multi-Granularity Cross-modal Alignment for Generalized Medical Visual Representation Learning. arXiv [cs.CV] (2022).

61. Santomauro, A., Portinale, L. & Leonardi, G. A multimodal approach to automated generation of radiology reports using contrastive learning (SHORT PAPER). 16–23 (2022).

62. Liang, G. et al. Contrastive Cross-Modal Pre-Training: A General Strategy for Small Sample Medical Imaging. IEEE J Biomed Health Inform 26, 1640–1649 (2022).

63. Zhang, Y., Jiang, H., Miura, Y., Manning, C. D. & Langlotz, C. P. Contrastive Learning of Medical Visual Representations from Paired Images and Text. arXiv [cs.CV] (2020).

64. Dadoun, H., Delingette, H., Rousseau, A.-L., Kerviler, E. & Ayache, N. Joint representation learning from french radiological reports and ultrasound images. Proc. IEEE Int. Symp. Biomed. Imaging 1–5 (2023).

65. Li, K. et al. DeAF: A multimodal deep learning framework for disease prediction. Comput. Biol. Med. 156, 106715 (2023).

66. Hadjiyski, N., Vosoughi, A. & Wismüller, A. Cross modal global local representation learning from radiology reports and x-ray chest images. in Medical Imaging 2023: Computer-Aided Diagnosis vol. 12465 722–731 (SPIE, 2023).

67. Pachade, S. et al. SELF-SUPERVISED LEARNING WITH RADIOLOGY REPORTS, A COMPARATIVE ANALYSIS OF STRATEGIES FOR LARGE VESSEL OCCLUSION AND BRAIN CTA IMAGES. Proc. IEEE Int. Symp. Biomed. Imaging 2023, (2023).

68. Zhang, S., et al. Large-Scale Domain-Specific Pretraining for Biomedical Vision-Language Processing. arXiv [cs.CV] (2023).

69. Bannur, S. et al. Learning to exploit temporal structure for biomedical vision-language processing. Proc. IEEE Comput. Soc. Conf. Comput. Vis. Pattern Recognit. 15016–15027 (2023).

70. Huang, Z., Bianchi, F., Yuksekgonul, M., Montine, T. J. & Zou, J. A visual–language foundation model for pathology image analysis using medical Twitter. Nat. Med. 29, 2307– 2316 (2023).

71. Lin, Z., Bas, E., Singh, K. Y., Swaminathan, G. & Bhotika, R. Relaxing contrastiveness in multimodal representation learning. Proc. IEEE Workshop Appl. Comput. Vis. 2226–2235 (2023).

72. Xue, Y. & Huang, X. Improved Disease Classification in Chest X-Rays with Transferred Features from Report Generation. in Information Processing in Medical Imaging 125–138 (Springer International Publishing, 2019).

73. Lee, C. et al. Classification of femur fracture in pelvic X-ray images using meta-learned deep neural network. Sci. Rep. 10, 13694 (2020).

74. Li, Y., Wang, H. & Luo, Y. A comparison of pre-trained vision-and-language models for multimodal representation learning across medical images and reports. Bioinform Biomed 1999–2004 (2020).

75. Wang, J. et al. MHKD-MVQA: Multimodal hierarchical knowledge distillation for medical Visual Question Answering. Bioinform Biomed 567–574 (2022).

76. Quigley, K., et al. RadTex: Learning Efficient Radiograph Representations from Text Reports. arXiv [cs.CV] (2022).

77. Li, C. et al. LLaVA-Med: Training a Large Language-and-Vision Assistant for Biomedicine in One Day. arXiv [cs.CV] (2023).

78. Kim, G., et al. KU-DMIS-MSRA at RadSum23: Pre-trained Vision-Language Model for Radiology Report Summarization. arXiv [cs.CL] (2023).

79. Moor, M., et al. Med-Flamingo: a Multimodal Medical Few-shot Learner. arXiv [cs.CV] (2023).

80. Chaves, J. M. Z. et al. Towards a clinically accessible radiology foundation model: open-access and lightweight, with automated evaluation. arXiv [cs.CL] (2024).

81. Saab, K., et al. Capabilities of Gemini Models in Medicine. arXiv [cs.AI] (2024).

82. Chen, Z., et al. CheXagent: Towards a Foundation Model for Chest X-Ray Interpretation. arXiv [cs.CV] (2024).

83. Tu, T., et al. Towards Generalist Biomedical AI. arXiv [cs.CL] (2023).

84. Zhou, H.-Y., Adithan, S., Acosta, J. N., Topol, E. J. & Rajpurkar, P. A Generalist Learner for Multifaceted Medical Image Interpretation. arXiv [cs.CV] (2024).

85. Hyland, S. L., et al. MAIRA-1: A specialised large multimodal model for radiology report generation. arXiv [cs.CL] (2023).

86. Bannur, S., et al. MAIRA-2: Grounded Radiology Report Generation. arXiv [cs.CL] (2024).

87. Chambon, P., et al. RoentGen: Vision-Language Foundation Model for Chest X-ray Generation. arXiv [cs.CV] (2022).

88. Chambon, P., Bluethgen, C., Langlotz, C. P. & Chaudhari, A. Adapting Pretrained Vision-Language Foundational Models to Medical Imaging Domains. arXiv [cs.CV] (2022).

89. Gu, Y., et al. BiomedJourney: Counterfactual Biomedical Image Generation by Instruction-Learning from Multimodal Patient Journeys. arXiv [cs.CV] (2023).

90. Nakaura, T. et al. The impact of large language models on radiology: a guide for radiologists on the latest innovations in AI. Jpn. J. Radiol. 42, 685–696 (2024).

91. Huang, S.-C. et al. Self-supervised learning for medical image classification: a systematic review and implementation guidelines. NPJ Digit Med 6, 74 (2023).

92. Liu, H. et al. M3AE: Multimodal Representation Learning for Brain Tumor Segmentation with Missing Modalities. AAAI 37, 1657–1665 (2023).

93. Touati, R. & Kadoury, S. A least square generative network based on invariant contrastive feature pair learning for multimodal MR image synthesis. Int. J. Comput. Assist. Radiol. Surg. 18, 971–979 (2023).

94. Fedorov, A., et al. On self-supervised multi-modal representation learning: An application to Alzheimer’s disease. in IEEE 18th International Symposium on Biomedical Imaging (2021).

95. Huang, R. et al. AW3M: An auto-weighting and recovery framework for breast cancer diagnosis using multi-modal ultrasound. Med. Image Anal. 72, 102137 (2021).

96. Liu, Y. et al. Incomplete multi-modal representation learning for Alzheimer’s disease diagnosis. Med. Image Anal. 69, 101953 (2021).

97. Fedorov, A. et al. Self-supervised multimodal learning for group inferences from MRI data: Discovering disorder-relevant brain regions and multimodal links. Neuroimage 285, 120485 (2024).

98. Meng, Z. et al. MSMFN: An Ultrasound Based Multi-Step Modality Fusion Network for Identifying the Histologic Subtypes of Metastatic Cervical Lymphadenopathy. IEEE Trans. Med. Imaging 42, 996–1008 (2023).

99. Cardoso, M. J., Sudre, C. H., Modat, M. & Ourselin, S. Template-Based Multimodal Joint Generative Model of Brain Data. Inf. Process. Med. Imaging 24, 17–29 (2015).

100. Huang, Y., Beltrachini, L., Shao, L. & Frangi, A. F. Geometry Regularized Joint Dictionary Learning for Cross-Modality Image Synthesis in Magnetic Resonance Imaging. in Simulation and Synthesis in Medical Imaging 118–126 (Springer International Publishing, 2016).

101. Yawen Huang, Ling Shao & Frangi, A. F. Cross-Modality Image Synthesis via Weakly Coupled and Geometry Co-Regularized Joint Dictionary Learning. IEEE Trans. Med. Imaging 37, 815–827 (2018).

102. Pan, Y. et al. Synthesizing Missing PET from MRI with Cycle-consistent Generative Adversarial Networks for Alzheimer’s Disease Diagnosis. Med. Image Comput. Comput. Assist. Interv. 11072, 455–463 (2018).

103. Peng, B., Huang, X., Wang, S. & Jiang, J. A REAL-TIME MEDICAL ULTRASOUND SIMULATOR BASED ON A GENERATIVE ADVERSARIAL NETWORK MODEL. Proc. Int. Conf. Image Proc. 2019, 4629–4633 (2019).

104. Wei, W. et al. Predicting PET-derived demyelination from multimodal MRI using sketcher-refiner adversarial training for multiple sclerosis. Med. Image Anal. 58, 101546 (2019).

105. Rana, A. et al. Use of Deep Learning to Develop and Analyze Computational Hematoxylin and Eosin Staining of Prostate Core Biopsy Images for Tumor Diagnosis. JAMA Netw Open 3, e205111 (2020).

106. Sharma, A. & Hamarneh, G. Missing MRI Pulse Sequence Synthesis Using Multi-Modal Generative Adversarial Network. IEEE Trans. Med. Imaging 39, 1170–1183 (2020).

107. Lan, H., Alzheimer Disease Neuroimaging Initiative, Toga, A. W. & Sepehrband, F. Three-dimensional self-attention conditional GAN with spectral normalization for multimodal neuroimaging synthesis. Magn. Reson. Med. 86, 1718–1733 (2021).

108. Raju, J., Murugesan, B., Ram, K. & Sivaprakasam, M. AutoSyncoder: An Adversarial AutoEncoder Framework for Multimodal MRI Synthesis. in Machine Learning for Medical Image Reconstruction 102–110 (Springer International Publishing, 2020).

109. Jiao, J., Namburete, A. I. L., Papageorghiou, A. T. & Noble, J. A. Self-Supervised Ultrasound to MRI Fetal Brain Image Synthesis. IEEE Trans. Med. Imaging 39, 4413–4424 (2020).

110. Morano, J., Hervella, Á. S., Barreira, N., Novo, J. & Rouco, J. Multimodal Transfer Learning-based Approaches for Retinal Vascular Segmentation. arXiv [eess.IV] (2020).

111. Dai, X. et al. Multimodal MRI synthesis using unified generative adversarial networks. Med. Phys. 47, 6343–6354 (2020).

112. Pan, Y., Liu, M., Lian, C., Xia, Y. & Shen, D. Spatially-Constrained Fisher Representation for Brain Disease Identification With Incomplete Multi-Modal Neuroimages. IEEE Trans. Med. Imaging 39, 2965–2975 (2020).

113. Zhan, B., Li, D., Wu, X., Zhou, J. & Wang, Y. Multi-Modal MRI Image Synthesis via GAN With Multi-Scale Gate Mergence. IEEE J Biomed Health Inform 26, 17–26 (2022).

114. Liu, H. et al. Improved amyloid burden quantification with nonspecific estimates using deep learning. Eur. J. Nucl. Med. Mol. Imaging 48, 1842–1853 (2021).

115. Zhang, L. et al. Spatial adaptive and transformer fusion network (STFNet) for low-count PET blind denoising with MRI. Med. Phys. 49, 343–356 (2022).

116. Lin, W. et al. Bidirectional Mapping of Brain MRI and PET With 3D Reversible GAN for the Diagnosis of Alzheimer’s Disease. Front. Neurosci. 15, 646013 (2021).

117. Fei, Y. et al. Deep learning-based multi-modal computing with feature disentanglement for MRI image synthesis. Med. Phys. 48, 3778–3789 (2021).

118. Hervella, Á. S., Rouco, J., Novo, J. & Ortega, M. Multimodal image encoding pre-training for diabetic retinopathy grading. Comput. Biol. Med. 143, 105302 (2022).

119. Fan, C., Lin, H. & Qiu, Y. U-Patch GAN: A Medical Image Fusion Method Based on GAN. J. Digit. Imaging 36, 339–355 (2023).

120. Sun, H. et al. Research on new treatment mode of radiotherapy based on pseudo-medical images. Comput. Methods Programs Biomed. 221, 106932 (2022).

121. Zhang, J., He, X., Qing, L., Gao, F. & Wang, B. BPGAN: Brain PET synthesis from MRI using generative adversarial network for multi-modal Alzheimer’s disease diagnosis. Comput. Methods Programs Biomed. 217, 106676 (2022).

122. Li, J. et al. TCGAN: a transformer-enhanced GAN for PET synthetic CT. Biomed. Opt. Express 13, 6003–6018 (2022).

123. Tang, L., Hui, Y., Yang, H., Zhao, Y. & Tian, C. Medical image fusion quality assessment based on conditional generative adversarial network. Front. Neurosci. 16, 986153 (2022).

124. Tawfik, N., Elnemr, H. A., Fakhr, M., Dessouky, M. I. & El-Samie, F. E. A. Multimodal Medical Image Fusion Using Stacked Auto-encoder in NSCT Domain. J. Digit. Imaging 35, 1308–1325 (2022).

125. Dalmaz, O., Yurt, M. & Cukur, T. ResViT: Residual Vision Transformers for Multimodal Medical Image Synthesis. IEEE Trans. Med. Imaging 41, 2598–2614 (2022).

126. Liu, S. & Yang, L. BPDGAN: A GAN-Based Unsupervised Back Project Dense Network for Multi-Modal Medical Image Fusion. Entropy 24, (2022).

127. Reaungamornrat, S., Sari, H., Catana, C. & Kamen, A. Multimodal image synthesis based on disentanglement representations of anatomical and modality specific features, learned using uncooperative relativistic GAN. Med. Image Anal. 80, 102514 (2022).

128. Wang, S., Zhang, X., Hui, H., Li, F. & Wu, Z. Multimodal CT Image Synthesis Using Unsupervised Deep Generative Adversarial Networks for Stroke Lesion Segmentation. Electronics 11, 2612 (2022).

129. Wang, A., Luo, X., Zhang, Z. & Wu, X.-J. A Disentangled Representation Based Brain Image Fusion via Group Lasso Penalty. Front. Neurosci. 16, 937861 (2022).

130. Liu, X. et al. CMRI2SPEC: CINE MRI SEQUENCE TO SPECTROGRAM SYNTHESIS VIA A PAIRWISE HETEROGENEOUS TRANSLATOR. Proc. IEEE Int. Conf. Acoust. Speech Signal Process. 2022, 1481–1485 (2022).

131. Cao, B. et al. AutoEncoder-Driven Multimodal Collaborative Learning for Medical Image Synthesis. Int. J. Comput. Vis. 131, 1995–2014 (2023).

132. Zhang, J., Zhang, S., Shen, X., Lukasiewicz, T. & Xu, Z. Multi-ConDoS: Multimodal Contrastive Domain Sharing Generative Adversarial Networks for Self-Supervised Medical Image Segmentation. IEEE Trans. Med. Imaging 43, 76–95 (2024).

133. Touati, R. & Kadoury, S. Bidirectional feature matching based on deep pairwise contrastive learning for multiparametric MRI image synthesis. Phys. Med. Biol. 68, (2023).

134. Li, W., Zhang, Y., Wang, G., Huang, Y. & Li, R. DFENet: A dual-branch feature enhanced network integrating transformers and convolutional feature learning for multimodal medical image fusion. Biomed. Signal Process. Control 80, 104402 (2023).

135. Xie, X. et al. MRSCFusion: Joint residual swin transformer and multiscale CNN for unsupervised multimodal medical image fusion. IEEE Trans. Instrum. Meas. 72, 1–17 (2023).

136. Taleb, A., Lippert, C., Klein, T. & Nabi, M. Multimodal Self-Supervised Learning for Medical Image Analysis. arXiv [cs.CV] (2019).

137. Xiang, Z. et al. Self-supervised multi-modal fusion network for multi-modal thyroid ultrasound image diagnosis. Comput. Biol. Med. 150, 106164 (2022).

138. van Tulder, G. & de Bruijne, M. Learning Cross-Modality Representations From Multi-Modal Images. IEEE Trans. Med. Imaging 38, 638–648 (2019).

139. Hou, Q., Peng, Y., Wang, Z., Wang, J. & Jiang, J. MFD-Net: Modality Fusion Diffractive Network for Segmentation of Multimodal Brain Tumor Image. IEEE J Biomed Health Inform 27, 5958–5969 (2023).

140. Zhou, Q. & Zou, H. A layer-wise fusion network incorporating self-supervised learning for multimodal MR image synthesis. Front. Genet. 13, 937042 (2022).

141. Fang, A. et al. Data Determines Distributional Robustness in Contrastive Language Image Pre-training (CLIP). in Proceedings of the 39th International Conference on Machine Learning (eds. Chaudhuri, K. et al.) vol. 162 6216–6234 (PMLR, 17--23 Jul 2022).

142. Huang, S.-C., et al. INSPECT: A multimodal dataset for patient outcome prediction of pulmonary embolisms. Adv. Neural Inf. Process. Syst. (2023).

143. Irvin, J., et al. CheXpert: A Large Chest Radiograph Dataset with Uncertainty Labels and Expert Comparison. arXiv [cs.CV] (2019).

144. Chambon, P., et al. CheXpert Plus: Augmenting a Large Chest X-ray Dataset with Text Radiology Reports, Patient Demographics and Additional Image Formats. arXiv [cs.CL] (2024).

145. Johnson, A. E. W., et al. MIMIC-CXR-JPG, a large publicly available database of labeled chest radiographs. arXiv [cs.CV] (2019).

146. Varma, M., Delbrouck, J.-B., Hooper, S., Chaudhari, A. & Langlotz, C. ViLLA: Fine-Grained Vision-Language Representation Learning from Real-World Data. in Proceedings of the IEEE/CVF International Conference on Computer Vision 22225–22235 (2023).

147. Boecking, B., et al. Making the Most of Text Semantics to Improve Biomedical Vision--Language Processing. arXiv [cs.CV] (2022).

148. Papineni, K., Roukos, S., Ward, T. & Zhu, W.-J. BLEU: a method for automatic evaluation of machine translation. in Proceedings of the 40th Annual Meeting on Association for Computational Linguistics 311–318 (Association for Computational Linguistics, USA, 2002).

149. Lin, C.-Y. ROUGE: A Package for Automatic Evaluation of Summaries. in Text Summarization Branches Out 74–81 (Association for Computational Linguistics, Barcelona, Spain, 2004).

150. Jain, S., et al. RadGraph: Extracting Clinical Entities and Relations from Radiology Reports. arXiv [cs.CL] (2021).

151. Chaves, J. Z., et al. RaLEs: A benchmark for Radiology Language Evaluations. Adv. Neural Inf. Process. Syst. (2023).

152. Van Veen, D. et al. Adapted large language models can outperform medical experts in clinical text summarization. Nat. Med. 30, 1134–1142 (2024).

153. Ostmeier, S., et al. GREEN: Generative Radiology Report Evaluation and Error Notation. arXiv [cs.CL] (2024).

154. Shah, N. H., Milstein, A. & Bagley PhD, S. C. Making Machine Learning Models Clinically Useful. JAMA 322, 1351–1352 (2019).

155. Obermeyer, Z., Powers, B., Vogeli, C. & Mullainathan, S. Dissecting racial bias in an algorithm used to manage the health of populations. Science 366, 447–453 (2019).

156. Seyyed-Kalantari, L., Liu, G., McDermott, M., Chen, I. Y. & Ghassemi, M. CheXclusion: Fairness gaps in deep chest X-ray classifiers. in Biocomputing 2021 232–243 (WORLD SCIENTIFIC, 2020).

157. Zhou, Y., et al. RadFusion: Benchmarking Performance and Fairness for Multimodal Pulmonary Embolism Detection from CT and EHR. arXiv [eess.IV] (2021).

158. Page, M. J. et al. The PRISMA 2020 statement: an updated guideline for reporting systematic reviews. Rev. Esp. Cardiol. 74, 790–799 (2021).

159. Liu, H., Li, C., Li, Y. & Lee, Y. J. Improved Baselines with Visual Instruction Tuning. arXiv [cs.CV] (2023).

160. Larson, D. B., Magnus, D. C., Lungren, M. P., Shah, N. H. & Langlotz, C. P. Ethics of Using and Sharing Clinical Imaging Data for Artificial Intelligence: A Proposed Framework. Radiology 295, 675–682 (2020).

161. Good machine learning practice for medical device development - Guiding Principles. International Medical Device Regulators Forum https://www.imdrf.org/consultations/good-machine-learning-practice-medical-device-development-guiding-principles.

