## Supplemental Materials for "Multimodal Foundation Models for Medical Imaging - A Systematic Review and Implementation Guidelines"

### 1. Full Extracted data

| **Study Type** | **Table Link** |
|---|---|
| Image & Non-Image Full Data Extraction | https://docs.google.com/spreadsheets/d/1KAuikeKjxh1O7z6dfdxRB8Mg1b2Q1oijSN8oyAFjFoE/edit?usp=sharing |
| Image & Image Full Data Extraction | https://docs.google.com/spreadsheets/d/1ZSPxzMIO-YhiilKsFgW2ZeArgd1GI_1ix5K8IE_ygsw/edit?usp=sharing |

### 2. Keyword groups

| **Group** | **Keywords** |
|---|---|
| AI | AI<br>Artificial Intelligence<br>Deep Learning<br>Machine learning |
| Self-supervised Learning / Fondation Models | Foundation Model*<br>Large Language Model*<br>Vision Language Model* |

| | |
|---|---|
| | Unified Vision Model*<br>LLM<br>VLM<br>UVM<br>representation learning<br>self-supervis*<br>self supervis*<br>contrastive learning<br>contrastive-learning<br>contrastive loss<br>contrastive-loss<br>contrastive training<br>contrastive-training<br>noise contrastive estimation<br>NCE<br>pretext learning<br>pretext-learning<br>pre-text learning<br>pretext task<br>pretext-task<br>pre-text task<br>autoencoder*<br>auto-encoder<br>auto encoder<br>diffusion model<br>generative model<br>generative ai |
| Medical Images | endoscop*<br>colonoscop*<br>gastroscop*<br>whole slide imaging<br>microscope slide, microscope slides<br>pathology imaging<br>pathology slid*<br>microscope<br>microscopy<br>ophthalmoscope<br>ophthalmoscopy<br>fundus photography<br>fundus imaging<br>retinal image<br>dermatoscope<br>dermatoscopy<br>medical imag*<br>diagnostic imag* |

| | |
|---|---|
| | radiolog* imag*<br>CT scan*<br>CT-scan*<br>computed tomography<br>computer-assisted tomography<br>CAT scan*<br>CAT-scan*<br>PET scan*<br>positron emission tomography<br>FDG-PET<br>PET-CT<br>PET/CT<br>MRI<br>magnetic resonance imaging<br>MR scan*<br>MR-scan*<br>MRI scan*<br>MRI-scan*<br>x-ray*<br>radiograph*<br>mammogram<br>mammography<br>fluoroscopy<br>ultrasound<br>ultrasonograph*<br>sonograph*<br>echocardio*<br>panorama*<br>spect-ct<br>spect scan<br>Scintigraphy<br>gamma scan<br>perfusion scan<br>Endoscopy<br>Colonoscopy<br>Gastroscopy<br>radiography<br>diagnostic imaging<br>tomography, x-ray computed<br>positron-emission tomography<br>Positron Emission Tomography Computed Tomography<br>Magnetic Resonance Imaging<br>Radiography<br>X-Rays<br>mammography<br>fluoroscopy<br>Ultrasonography<br>Echocardiography<br>Single Photon Emission Computed |

| | |
|---|---|
| | Tomography Computed Tomography<br>radionuclide imaging<br>Ventilation-Perfusion Scan |
| Non-imaging Modalities | genomic*<br>gene<br>genes<br>snp*<br>single nucleotide polymorphism<br>proteomic*<br>dna<br>rna<br>Multimodal<br>Health records<br>Medical records<br>Medication<br>Vitals<br>Lab test<br>radiology report<br>clinical note<br>physician* note<br>doctor* note |

### 3. Full Search String

| **Database** | **Search Strings** |
|---|---|
| Scopus | ((TITLE-ABS ("Foundation Model*") OR TITLE-ABS ("Large Language Model*") OR TITLE-ABS ("Vision Language Model*") OR TITLE-ABS ("Unified Vision Model*") OR TITLE-ABS ("LLM") OR TITLE-ABS ("VLM") OR TITLE-ABS ("UVM") OR TITLE-ABS ("representation learning") OR TITLE-ABS ("self-supervis*") OR TITLE-ABS ("self supervis*") OR TITLE-ABS ("contrastive learning") OR TITLE-ABS ("contrastive-learning") OR TITLE-ABS ("contrastive loss") OR TITLE-ABS ("contrastive-loss") OR TITLE-ABS ("contrastive training") OR TITLE-ABS ("contrastive-training") OR TITLE-ABS ("noise contrastive estimation") OR TITLE-ABS ("NCE") OR TITLE-ABS ("pretext learning") OR TITLE-ABS ("pretext-learning") OR TITLE-ABS ("pre-text learning") OR TITLE-ABS ("pretext task") OR TITLE-ABS ("pretext-task") OR TITLE-ABS ("pre-text task") OR TITLE-ABS ("autoencoder*") OR TITLE-ABS ("auto-encoder") OR TITLE-ABS ("auto encoder") OR TITLE-ABS ("diffusion model") OR TITLE-ABS ("generative model") OR TITLE-ABS ("generative ai")) AND (TITLE-ABS ("endoscop*") OR TITLE-ABS ("colonoscop*") OR TITLE-ABS ("gastroscop*") OR TITLE-ABS ("whole slide imaging") OR TITLE-ABS ("microscope slide, microscope slides") OR TITLE-ABS ("pathology imaging") |

| | |
|---|---|
| | OR TITLE-ABS ("pathology slid*") OR TITLE-ABS ("microscope") OR TITLE-ABS ("microscopy") OR TITLE-ABS ("ophthalmoscope") OR TITLE-ABS ("ophthalmoscopy") OR TITLE-ABS ("fundus photography") OR TITLE-ABS ("fundus imaging") OR TITLE-ABS ("retinal image") OR TITLE-ABS ("dermatoscope") OR TITLE-ABS ("dermatoscopy") OR TITLE-ABS ("medical imag*") OR TITLE-ABS ("diagnostic imag*") OR TITLE-ABS ("radiolog* imag*") OR TITLE-ABS ("CT scan*") OR TITLE-ABS ("CT-scan*") OR TITLE-ABS ("computed tomography") OR TITLE-ABS ("computer-assisted tomography") OR TITLE-ABS ("CAT scan*") OR TITLE-ABS ("CAT-scan*") OR TITLE-ABS ("PET scan*") OR TITLE-ABS ("positron emission tomography") OR TITLE-ABS ("FDG-PET") OR TITLE-ABS ("PET-CT") OR TITLE-ABS ("PET/CT") OR TITLE-ABS ("MRI") OR TITLE-ABS ("magnetic resonance imaging") OR TITLE-ABS ("MR scan*") OR TITLE-ABS ("MR-scan*") OR TITLE-ABS ("MRI scan*") OR TITLE-ABS ("MRI-scan*") OR TITLE-ABS ("x-ray*") OR TITLE-ABS ("radiograph*") OR TITLE-ABS ("mammogram") OR TITLE-ABS ("mammography") OR TITLE-ABS ("fluoroscopy") OR TITLE-ABS ("ultrasound") OR TITLE-ABS ("ultrasonograph*") OR TITLE-ABS ("sonograph*") OR TITLE-ABS ("echocardio*") OR TITLE-ABS ("panorama*") OR TITLE-ABS ("spect-ct") OR TITLE-ABS ("spect scan") OR TITLE-ABS ("scintigraphy") OR TITLE-ABS ("gamma scan") OR TITLE-ABS ("perfusion scan") OR TITLE-ABS ("Endoscopy") OR TITLE-ABS ("Colonoscopy") OR TITLE-ABS ("Gastroscopy") OR TITLE-ABS ("radiography") OR TITLE-ABS ("diagnostic imaging") OR TITLE-ABS ("tomography, x-ray computed") OR TITLE-ABS ("positron-emission tomography") OR TITLE-ABS ("Positron Emission Tomography Computed Tomography") OR TITLE-ABS ("Magnetic Resonance Imaging") OR TITLE-ABS ("Radiography") OR TITLE-ABS ("X-Rays") OR TITLE-ABS ("mammography") OR TITLE-ABS ("fluoroscopy") OR TITLE-ABS ("Ultrasonography") OR TITLE-ABS ("Echocardiography") OR TITLE-ABS ("Single Photon Emission Computed Tomography Computed Tomography") OR TITLE-ABS ("radionuclide imaging") OR TITLE-ABS ("Ventilation-Perfusion Scan")) AND (TITLE-ABS ("genomic*") OR TITLE-ABS ("gene") OR TITLE-ABS ("genes") OR TITLE-ABS ("snp*") OR TITLE-ABS ("single nucleotide polymorphism ") OR TITLE-ABS ("proteomic*") OR TITLE-ABS ("dna") OR TITLE-ABS ("rna") OR TITLE-ABS ("Multimodal") OR TITLE-ABS ("Health records") OR TITLE-ABS ("Medical records") OR TITLE-ABS ("Medication") OR TITLE-ABS ("Vitals") OR TITLE-ABS ("Lab test") OR TITLE-ABS ("radiology report") OR TITLE-ABS ("clinical note") OR TITLE-ABS ("physician* note") OR TITLE-ABS ("doctor* note")) AND (LIMIT-TO ( PUBYEAR , 2023 ) OR LIMIT-TO ( PUBYEAR , 2022 ) OR LIMIT-TO ( PUBYEAR , 2021 ) OR LIMIT-TO ( PUBYEAR , 2020 ) OR LIMIT-TO ( PUBYEAR , 2019 ) OR LIMIT-TO ( PUBYEAR , 2018 ) OR LIMIT-TO ( PUBYEAR , 2017 ) OR LIMIT-TO ( PUBYEAR , 2016 ) OR LIMIT-TO ( PUBYEAR , 2015 ) OR LIMIT-TO ( PUBYEAR , 2014 ) OR LIMIT-TO ( PUBYEAR , 2013 ) OR LIMIT-TO ( PUBYEAR , 2012 ))) |
| Pubmed | ("Foundation Model*"[Title/Abstract] OR "Large Language Model*"[Title/Abstract] OR "Vision Language Model*"[Title/Abstract] OR "Unified Vision Model*"[Title/Abstract] OR "LLM"[Title/Abstract] OR "VLM"[Title/Abstract] OR "UVM"[Title/Abstract] OR "representation learning"[Title/Abstract] OR |

| | |
|---|---|
| | "self-supervis*"[Title/Abstract] OR "self supervis*"[Title/Abstract] OR "contrastive learning"[Title/Abstract] OR "contrastive-learning"[Title/Abstract] OR "contrastive loss"[Title/Abstract] OR "contrastive-loss"[Title/Abstract] OR "contrastive training"[Title/Abstract] OR "contrastive-training"[Title/Abstract] OR "noise contrastive estimation"[Title/Abstract] OR "NCE"[Title/Abstract] OR "pretext learning"[Title/Abstract] OR "pretext-learning"[Title/Abstract] OR "pre-text learning"[Title/Abstract] OR "pretext task"[Title/Abstract] OR "pretext-task"[Title/Abstract] OR "pre-text task"[Title/Abstract] OR "autoencoder*"[Title/Abstract] OR "auto-encoder"[Title/Abstract] OR "auto encoder"[Title/Abstract] OR "diffusion model"[Title/Abstract] OR "generative model"[Title/Abstract] OR "generative ai"[Title/Abstract]) AND ("endoscop*"[Title/Abstract] OR "colonoscop*"[Title/Abstract] OR "gastroscop*"[Title/Abstract] OR "whole slide imaging"[Title/Abstract] OR "microscope slide, microscope slides"[Title/Abstract] OR "pathology imaging"[Title/Abstract] OR "pathology slid*"[Title/Abstract] OR "microscope"[Title/Abstract] OR "microscopy"[Title/Abstract] OR "ophthalmoscope"[Title/Abstract] OR "ophthalmoscopy"[Title/Abstract] OR "fundus photography"[Title/Abstract] OR "fundus imaging"[Title/Abstract] OR "retinal image"[Title/Abstract] OR "dermatoscope"[Title/Abstract] OR "dermatoscopy"[Title/Abstract] OR "medical imag*"[Title/Abstract] OR "diagnostic imag*"[Title/Abstract] OR "radiolog* imag*"[Title/Abstract] OR "CT scan*"[Title/Abstract] OR "CT-scan*"[Title/Abstract] OR "computed tomography"[Title/Abstract] OR "computer-assisted tomography"[Title/Abstract] OR "CAT scan*"[Title/Abstract] OR "CAT-scan*"[Title/Abstract] OR "PET scan*"[Title/Abstract] OR "positron emission tomography"[Title/Abstract] OR "FDG-PET"[Title/Abstract] OR "PET-CT"[Title/Abstract] OR "PET/CT"[Title/Abstract] OR "MRI"[Title/Abstract] OR "magnetic resonance imaging"[Title/Abstract] OR "MR scan*"[Title/Abstract] OR "MR-scan*"[Title/Abstract] OR "MRI scan*"[Title/Abstract] OR "MRI-scan*"[Title/Abstract] OR "x-ray*"[Title/Abstract] OR "radiograph*"[Title/Abstract] OR "mammogram"[Title/Abstract] OR "mammography"[Title/Abstract] OR "fluoroscopy"[Title/Abstract] OR "ultrasound"[Title/Abstract] OR "ultrasonograph*"[Title/Abstract] OR "sonograph*"[Title/Abstract] OR "echocardio*"[Title/Abstract] OR "panorama*"[Title/Abstract] OR "spect-ct"[Title/Abstract] OR "spect scan"[Title/Abstract] OR "Scintigraphy"[Title/Abstract] OR "gamma scan"[Title/Abstract] OR "perfusion scan"[Title/Abstract] OR "Endoscopy"[MeSH Terms] OR "Colonoscopy"[MeSH Terms] OR "Gastroscopy"[MeSH Terms] OR "radiography"[MeSH Terms] OR "diagnostic imaging"[MeSH Terms] OR "tomography, x-ray computed"[MeSH Terms] OR "positron-emission tomography"[MeSH Terms] OR "Positron Emission Tomography Computed Tomography"[MeSH Terms] OR "Magnetic Resonance Imaging"[MeSH Terms] OR "Radiography"[MeSH Terms] OR "X-Rays"[MeSH Terms] OR "mammography"[MeSH Terms] OR "fluoroscopy"[MeSH Terms] OR "Ultrasonography"[MeSH Terms] OR "Echocardiography"[MeSH Terms] OR "Single Photon Emission Computed Tomography Computed Tomography"[MeSH Terms] OR "radionuclide imaging"[MeSH Terms] OR "Ventilation-Perfusion Scan"[MeSH Terms]) AND ("genomic*"[Title/Abstract] OR |

| | |
|---|---|
| | "gene"[Title/Abstract] OR "genes"[Title/Abstract] OR "snp*"[Title/Abstract] OR "single nucleotide polymorphism "[Title/Abstract] OR "proteomic*"[Title/Abstract] OR "dna"[Title/Abstract] OR "rna"[Title/Abstract] OR "Multimodal"[Title/Abstract] OR "Health records"[Title/Abstract] OR "Medical records"[Title/Abstract] OR "Medication"[Title/Abstract] OR "Vitals"[Title/Abstract] OR "Lab test"[Title/Abstract] OR "radiology report"[Title/Abstract] OR "clinical note"[Title/Abstract] OR "physician* note"[Title/Abstract] OR "doctor* note"[Title/Abstract] OR "Genomics"[MeSH Terms] OR "Genome"[MeSH Terms] OR "Genes"[MeSH Terms] OR "Polymorphism, Single Nucleotide"[MeSH Terms] OR "Proteomics"[MeSH Terms] OR "DNA"[MeSH Terms] OR "RNA"[MeSH Terms]) AND ("2012/01/01"[PDAT] : "2024/01/01"[PDAT]) |
